# Early prediction of ovarian cancer risk based on real world data

**DOI:** 10.1101/2024.07.26.24310994

**Authors:** Víctor de la Oliva, Alberto Esteban-Medina, Laura Alejos, Dolores Muñoyerro-Muñiz, Román Villegas, Joaquín Dopazo, Carlos Loucera

## Abstract

This study presents the development of an early prediction model for high-grade serous ovarian cancer (HGSOC) using real-world data from the Andalusian Health Population Database (BPS), containing electronic health records (EHR) of over 15 million patients. Leveraging the extensive data availability, the model aims to identify individuals at high risk of HGSOC without the need for specific tumor markers or prior stratification into risk groups. Utilizing an Explainable Boosting Machine (EBM) algorithm, the model incorporates diverse clinical variables including demographics, chronic diseases, symptoms, blood test results, and healthcare utilization patterns. The model was trained and validated using a total of 3,088 HGSOC patients diagnosed between 2018 and 2022 along with 114,942 controls of similar characteristics, to emulate the prevalence of the disease, achieving a sensitivity of 0.65 and a specificity of 0.85. This study underscores the importance of using patient data from the general population, demonstrating that effective early detection models can be developed from routinely collected healthcare data. The approach addresses limitations of traditional screening methods by providing a cost-effective and broadly applicable tool for early cancer detection, potentially improving patient outcomes through timely interventions. The interpretability of the early prediction model also offers insights into the most significant predictors of cancer risk, further enhancing its utility in clinical settings.

## Introduction

Ovarian cancer (OC) is considered one of the most serious tumors in women and is the leading cause of death among all gynecological tumors [1], with an incidence of approximately 1 in 78 [2]. Paradoxically, despite its high mortality rate (over 75%), its cure rate when diagnosed early (when the tumor is still confined to the ovaries, FIGO stage I) is very high (90%) [3]. However, delays in diagnosis are very common in this disease, and are usually due to unawareness of the initial symptoms of ovarian cancer by patients, lack of information in primary care, or delays in requesting accurate diagnostic tests. As a consequence of these delays, at the time of diagnosis, many OC patients present an advanced stage of the disease (the tumor has spread out of the ovaries and has taken other nearby structures) and the recommended interventions to treat ovarian cancer, such as surgery or chemotherapy, become no longer an option leaving palliative care as the only alternative [4]. Epithelial cancers are the most common OCs, accounting for 90% of all cases, and among these, 58% are serous [5], being the vast majority of them HGSOC [6].

Because its prevalence is low in the general population (about 600 cases are detected each year in Andalusia), any screening strategy should not only be very sensitive in the initial stages, but also highly specific [7]. Some attempts to develop effective ovarian cancer screening strategies have used traditional clinical interventions such as transvaginal ultrasound, or serum tumor markers, such as CA125 [8]. However, none of these methods, when used occasionally and individually, present an adequate predictive value and, on the contrary, their systematic use is expensive [4]. Although new markers with the potential to be used in diagnosis are being detected with the developments of genomics and proteomics methodologies, they still require prospective validation work regarding their specificity and sensitivity, and the truth is that, to date, despite the clear benefit of early diagnosis for patients, there is no strategy for screening and early detection of HGSOC risk.

Another strategy to help in potential screenings is the use of end-point predictors based on clinical real-world data (RWD). Actually, Real World Evidence (RWE) studies on retrospective cohorts have resulted in quite accurate predictions of the evolution of the disease in distinct patient types. For example, *Deep Patient* allows predicting the development of various diseases with 90% accuracy [9], *Doctor AI*, makes preventive future diagnoses and recommends treatments [10] or *Deepcare*, which predicts disease progression, recommends interventions and estimates future risks [11] are examples of successful application of Machine Learning techniques to large clinical data repositories. Such strategies do not require a previous patient stratification into a risk group and can be used in the general population as a pre-screening strategy. A recent publication illustrates this concept in OC, although the number of samples in the database used, the Surveillance, Epidemiology, and End Results (SEER) dataset, is reduced, which casts doubts on the reproducibility of the results when applied to a broader population [12]. Similar problems due to a low training dataset occur in other works [13,14].

Andalusia, the third largest region in Europe, with 8.5 million inhabitants [15], a population similar to medium-sized European countries like Austria or Switzerland, has a universal electronic health record (EHR) managed by an efficient digital system called Diraya. Patient’s information in Diraya is systematically uploaded to the Population Health Base (BPS) on a monthly basis. This makes BPS one of the largest repositories of highly detailed clinical data in the world (with over 15 million patients) [16]. To facilitate the secondary use of biomedical data for clinical research purposes, a trusted research environment (TRE) [17], the Infrastructure for Secure generation of Evidence from Real World Data from the Population Health Database of Andalusia (iRWD) [18], was recently implemented to allow secure analysis of clinical data. Notable recent initiatives utilizing extensive real-world data from BPS include investigations into the protective role vitamin D in COVID-19 patients [19], research subsequently expanded on other drugs [20], the impact of oral anticoagulants on atrial fibrillation-related stroke [21], or an analysis of how different SARS-CoV-2 variants impact COVID-19 patient survival [22], just to cite a few ones.

Hence, BPS offers a unique environment for a thorough retrospective examination of ovarian cancer patient profiles, yielding real-world evidence on healthcare system utilization patterns. The objective of this study is to create a machine learning predictor for early detection of high-grade serous ovarian cancer (HGSOC) using uniquely data from the electronic health records available in the BPS database.

## Material and Methods

### Data source

The Ethics Committee for the Coordination of Biomedical Research in Andalusia granted approval for the study titled “Retrospective observational study for the development of early predictor of ovarian cancer” (29th March, 2022, Acta 03/22) and waived informed consent for the secondary use of clinical data for research purposes. Real world longitudinal data sets were extracted from the BPS [16]. These datasets are derived from patient EHRs from the Andalusian public health system (i.e. all data from users of the Andalusian public health system). In addition to clinical data, these datasets also contain patient demographic data such as age, district of residence or socioeconomic data and represent virtually the entire population in the Andalusian region. Since BPS contains data from the whole Andalusian Health System since 2017, the study period was 2018 to 2022, allowing thus the availability of, at least, one previous year of information for all the individuals studied.

A total of 3,088 patients diagnosed with ovarian cancer (OC) in Andalusia from 2018 to 2022, aged 50 or older at diagnosis time, were extracted from BPS. The distribution of cases across the years is as follows: 572 diagnosed in 2018, 586 in 2019, 607 in 2020, 631 in 2021, and 692 in 2022. In order to approximately mimic the prevalence of the disease, a total of 114,942 controls, matching the age and district of residence of the cases, were also extracted from the database.

### Variables used

In this study, different variables with known or suspected impact in risk of developing OC are collected that include: i) Sociodemographic data (sex, birth date, death date, district); ii) Chronic diseases, preprocessed by BPS curators from ICD10 codes and free text [16]. BPS pathologies have diagnosis date and have been grouped into disease categories (see Table 1); iii) Symptoms and diagnoses of non-chronic illnesses categorized by their ICD10 code, selected by BPS curators and the OC literature [23–25] (Supplementary table S1). The occurrence of these during the observation window was treated as a binary variable; iv) Analytic tests consisting of ordinary clinical petitions (biochemistry parameters in blood and urine analyses). Scientific action protocols were revised [26] to check the inclusion of other parameters (Supplementary Table S2); vi) Number of visits to relevant medical specialties (gynecology and gastroenterology) during the observation window (Supplementary Table S3).

**Table 1:** Overall and case-control stratified counts and proportions of comorbidities associated to the study cohort with p-values corresponding to case-control *X*^2^/ANOVA tests. For cases, these are comorbidities diagnosed before their ovarian cancer diagnosis and for controls, they are those diagnosed before a randomly attributed index point matching the diagnosis date of ovarian cancer patients.

| Comorbidity | Overall | Case | Control | P-Value |
| --- | --- | --- | --- | --- |
| **Age-Related Macular Degeneration, n (%)** | 2299 (1.9) | 47 (1.5) | 2252 (2.0) | 0.095 |
| **Arthritis And Arthrosis, n (%)** | 57635 (48.8) | 1392 (45.1) | 56243 (48.9) | <0.001 |
| **Bladder Cancer, n (%)** | 403 (0.3) | 24 (0.8) | 379 (0.3) | <0.001 |
| **Blood Cancer, n (%)** | 898 (0.8) | 29 (0.9) | 869 (0.8) | 0.293 |
| **Bone and Soft Tissue Cancer, n (%)** | 167 (0.1) | 30 (1.0) | 137 (0.1) | <0.001 |
| **Breast Cancer, n (%)** | 4719 (4.0) | 185 (6.0) | 4534 (3.9) | <0.001 |
| **Bronchus and Lung Cancer, n (%)** | 364 (0.3) | 29 (0.9) | 335 (0.3) | <0.001 |
| **Cardiovascular Disease, n (%)** | 65177 (55.2) | 1679 (54.4) | 63498 (55.2) | 0.346 |
| **Digestive System Cancer, n (%)** | 2246 (1.9) | 131 (4.2) | 2115 (1.8) | <0.001 |
| **Digestive System Disease, n (%)** | 2867 (2.4) | 76 (2.5) | 2791 (2.4) | 0.954 |
| **Endocrine Disease, n (%)** | 71946 (61.0) | 1898 (61.5) | 70048 (60.9) | 0.57 |
| **Eye Disease, n (%)** | 11390 (9.7) | 265 (8.6) | 11125 (9.7) | 0.045 |
| **Gastrointestinal Disease, n (%)** | 7684 (6.5) | 238 (7.7) | 7446 (6.5) | 0.007 |
| **Genitourinary Cancer, n (%)** | 309 (0.3) | 19 (0.6) | 290 (0.3) | <0.001 |
| **Genitourinary Disease, n (%)** | 15404 (13.1) | 500 (16.2) | 14904 (13.0) | <0.001 |
| **Gynecological Cancers, n (%)** | 1355 (1.1) | 186 (6.0) | 1169 (1.0) | <0.001 |
| **Head and Neck Cancer, n (%)** | 296 (0.3) | 2 (0.1) | 294 (0.3) | 0.056 |
| **Infectious Disease, n (%)** | 152 (0.1) | 4 (0.1) | 148 (0.1) | 1 |
| **Kaposi's Sarcoma, n (%)** | 8 (0.0) | 0 | 8 (0.0) | 1 |
| **Liver Disease, n (%)** | 7835 (6.6) | 280 (9.1) | 7555 (6.6) | <0.001 |
| **Melanoma, n (%)** | 471 (0.4) | 12 (0.4) | 459 (0.4) | 1 |
| **Mental Illness, n (%)** | 50342 (42.7) | 1399 (45.3) | 48943 (42.6) | 0.003 |
| **Musculoskeletal Condition, n (%)** | 24584 (20.8) | 581 (18.8) | 24003 (20.9) | 0.006 |
| **Nervous System Disease, n (%)** | 8844 (7.5) | 201 (6.5) | 8643 (7.5) | 0.038 |
| **Respiratory Disease, n (%)** | 15499 (13.1) | 408 (13.2) | 15091 (13.1) | 0.914 |
| **Skin Disease, n (%)** | 3462 (2.9) | 97 (3.1) | 3365 (2.9) | 0.522 |
| **Thyroid Cancer, n (%)** | 413 (0.3) | 24 (0.8) | 389 (0.3) | <0.001 |
| **Total N** | 118030 | 3088 | 114942 |  |
| **Age, median [Q1, Q3]** | 68.0 [59.0, 78.0] | 64.0 [57.0, 74.0] | 68.0 [59.0, 78.0] | <0.001 |

Highly sparse variables, like hormonal biochemistry, genito-urinary system-related pathogenic culture results, STD blood tests, or variables directly related to OC suspicion, as tumor marker determinations, were removed from the study.

### Modeling

#### Training, validation, and test sets

The OC patients were stratified based on their diagnosis dates for different phases of the model development: training (2018-2020, N=1,765), validation (2021, N=631), and testing (2022, N=692).

The control non-OC patients were divided into three random disjoint sets: 40% for training, 30% for validation, and the remaining 30% for testing. The test group was never used to train or to perform model selection.

#### Modeling methods

The model aims to discriminate between cases, defined as women with an ovarian cancer (OC) diagnosis, and controls, which consisted of women who exhibited similar demographic and health profiles but lacked an OC diagnosis. The objective was to discriminate at the earliest possible stage using longitudinal patient data. To achieve this, a time dimension was incorporated into the model during training. For each individual, an “index point” was established, corresponding to the diagnosis date for cases and a randomly selected date from within the timeframe of the training dataset for controls. Subsequently, raw patient data was transformed to facilitate analysis. These transformations involved the grouping of BPS chronic comorbidities (Table 1), conversion of symptoms and acute illnesses into binary variables, and the aggregation of data pertaining to medical specialty visits and blood test results over a predetermined observation window. The sole demographic variable incorporated was the age of the patient at the time of evaluation.

For training purposes, data within a predefined *leave-out window* immediately preceding the index point, also known as the horizon, was excluded. The model was then trained on the remaining data captured within a specified *feature window*, termed the history, which preceded the *leave-out window*. An Explainable Boosting Machine (EBM) algorithm [27] was used for its capacity to provide transparent explanations of its predictive decisions. The model employed to fit the data was designated as the *base estimator*. This model represents a specialized form of the generalized additive model (GAM) [28], wherein tree-based boosting methods are employed to learn the shape functions [29]. The fitting pipeline is specifically designed to enhance the interpretability of both the individual shape functions and the model as a whole.

The EBM algorithm constructs each shape function through an ensemble of gradient-boosted trees (GBTs). This process involves the sequential, iterative handling of all features. For each feature, a shallow tree, characterized by limited depth, is constructed using only that feature alongside a subset of the data. Concurrently, residuals are updated following a boosting methodology. As a result, each tree is restricted to the feature it was trained on, thereby enabling precise learning of each feature’s contribution while simultaneously maintaining a global approximation via the residuals. This iterative process is repeated thousands of times, cycling through the data with a minimal learning rate, rendering the sequence of feature processing inconsequential. Upon completing all iterations, the model constructs a shape function by aggregating all the trees trained for any given feature. These shape functions are then collectively combined to establish the final decision rule of the model.

EBMs are considered a valuable tool for tasks where understanding the rationale behind predictions is as important as the predictions themselves, particularly in high-stakes scenarios such as healthcare or biomedicine, while also providing state-of-the-art performance [30]. The standard array of hyper-parameters is used to fit all EBMs, with the exception of bootstrap sampling rounds, which are increased as recommended [31].

To mirror continuous evaluation in real-world settings, a sequential prediction strategy was implemented during the validation and testing phases. Using this approach, the model generated monthly predictions for ovarian cancer (OC) diagnosis probability, utilizing only the clinical history data available up to each respective month. Specifically, for the validation set, which included patients diagnosed with OC in 2021, this method was applied monthly from 2018 through 2021. This approach produced a time series of sequential probability predictions for each patient, termed the probability trajectory. Furthermore, a decision threshold was determined based on the validation set to achieve a sensitivity of 58%. This sensitivity level was chosen to align with that reported in a prior ovarian cancer screening study that also accounted for non-compliant patients [32].

For patients who were eventually diagnosed with OC, the earliness of the model’s predictions can be quantified if, in any given month, their predicted probability of developing OC exceeds the learned threshold. However, it should be noted that these predictions are only meaningful if they occur before the actual date of confirmed diagnosis. Post-diagnosis evaluations for OC patients are discontinued, meaning that any instance where an OC patient’s probability trajectory does not exceed the threshold prior to diagnosis is considered a missed prediction by the model. Similarly, predictions indicating that non-OC patients are at risk of developing OC at any point are also classified as missed predictions. This control group is monitored throughout the entire duration of the study period.

#### Variable codification

Chronic diseases, symptoms and diagnoses are modeled as binary variables within a specified time window. If a diagnosis for a condition is received by a patient before the end of the historical window, the corresponding feature value is set to 1; otherwise, it is set to 0. Specialist visits to relevant medical specialties, such as gynecology and gastroenterology, are modeled as continuous variables, representing the total number of visits by each patient across the historical window. Regarding analytics, valid values are log-transformed while invalid values for each analytic are set to missing. Descriptive statistics, including mean, standard deviation, maximum, and minimum, are computed for each feature over the historical window. If a statistic is missing for a patient, the missing value is imputed using the population median calculated during the training phase. Finally, the age of the patient is recorded as of the end of the historical window.

### Evaluation

To evaluate the patients throughout time the following metrics are used:

- Recall (sensitivity): True Positives/(True Positives + False Negatives)
- Specificity: True Negatives /(False Positives + True Negatives)
- Precision: True Positives/(True Positives + False Positives)
- False positive rate: False Positive /(True Positive + False Positive)
- F1-score: 2*(Precision * Recall)/(Precision + Recall)
- Normalized earliness: (Days to diagnosis)/365

To facilitate the interpretation of the metrics, results are presented based on the split criterion. Initially, the metrics for the completely independent test partition are reported. Subsequently, the metrics for the full cohort (excluding the ovarian cancer cases used for training and validation) are reported to simulate the continuous evaluation of patients over time.

To evaluate the model’s performance before setting a threshold, the Area under the Receiver Operating Characteristic Curve (AUROC) and the Area under the Precision and Recall Curve (AUPRC) are reported, along with graphics depicting the corresponding curves. Additionally, the Area under the Precision-Recall-Gain Curve (AUPRGC) is measured [33]. Similar to the PRC, the PRG curve plots precision on the y-axis and recall on the x-axis, but it uses gain instead of raw precision values. Gain represents the difference between the current precision and a baseline value, reflecting a prevalence-informed random classification. This method is particularly useful for model selection when compared to a weak baseline, such as in cases with low prevalence. A higher PRG curve or AUPRG indicates better model performance.

Finally, the hyperbolic-weighted tau (hwt) statistic [34] is used to measure the correlation among rankings. This metric provides a balanced approach by reducing the impact of less informative ranking segments (the tail) while penalizing variations in the more informative segments (the head of the ranking). This method is a modified version of Kendall’s tau [35], where the correlation between two rankings is adjusted with an additive hyperbolic function that imposes greater penalties on discrepancies at the higher end of the rank. This is particularly suitable for examining disparities among the learned characteristics of models, as it penalizes inconsistencies where the models are more focused.

### Model selection

The optimal lengths for the history and horizon windows are determined by evaluating a model over the validation set through a grid search over a one-year period in both past and future directions, with window sizes evaluated in 30-day increments.

## Results

### Model selection

Panel A of Figure 1 displays the model’s AUPRC on the validation set for various combinations of prediction time windows (horizon) and historical data lengths used for training (history). The 30-day horizon window band shows consistent performance across all history window lengths and outperforms other horizon windows.

**Figure 1:**
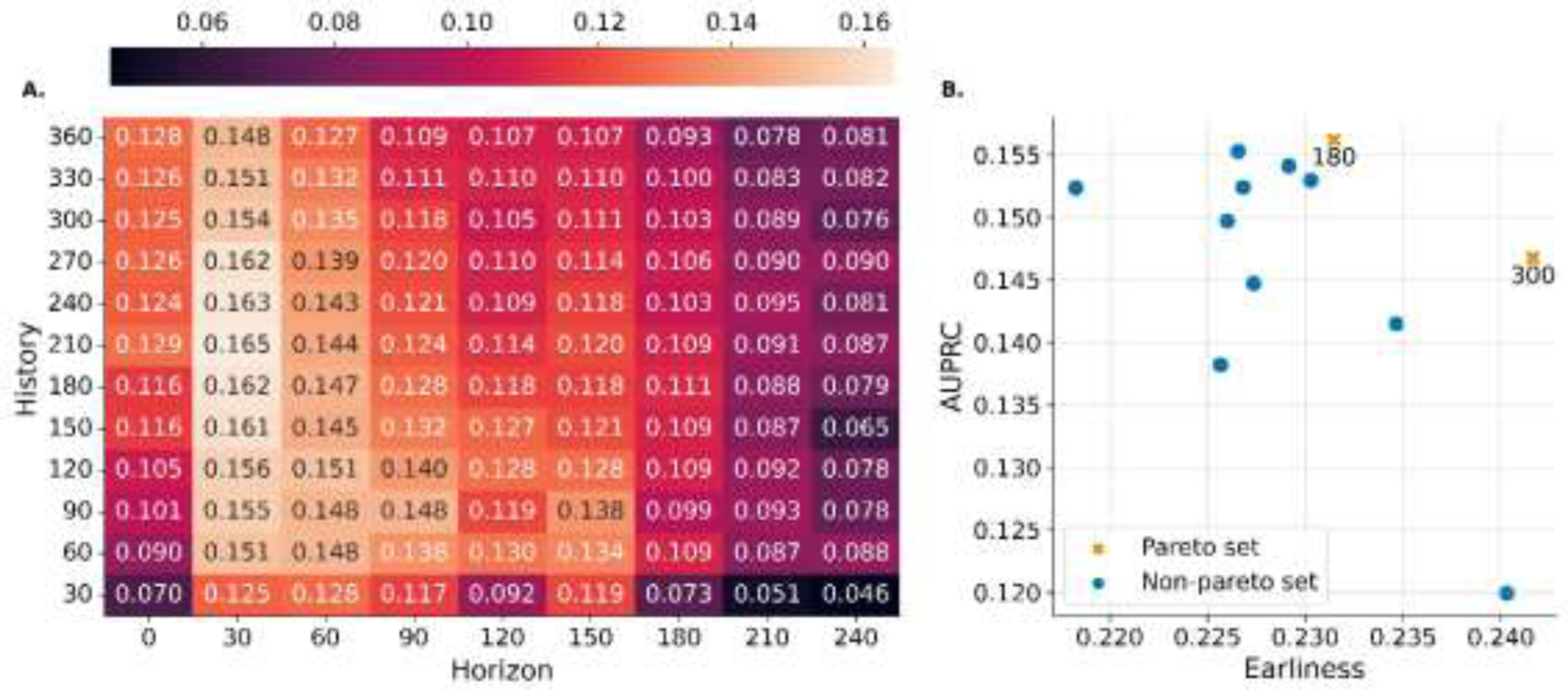
Performance summary across the validation set. A: Area Under the Precision-Recall Curve (AUPRC) on the validation set for various prediction horizons (x-axis) and training history lengths (y-axis). The 30-day horizon band shows consistent performance across history lengths and outperforms other horizons. B: Pareto set for the scatter plot of means, considering earliness (timeliness of prediction) vs. AUPRC. The selected model with a 180-day history window is highlighted, offering a balance between performance and potentially less data imputation.

To select a model from those trained with a 30-day horizon, the experiment was repeated using 100 train/validation splits, following the previously mentioned proportion rules and fixing the horizon window at 30 days. Panel B of Figure 1 presents the Pareto set for the scatter plot of means, considering earliness versus AUPRC. From the two models on the Pareto frontier, the model trained with a 180-day history window was selected. This choice potentially requires less imputation (due to its shorter window span) compared to models trained with 300-day history windows.

Therefore, a 30-day prediction horizon and a 180-day history window will be used for training. The full set of combinations tested, along with visualizations of each metrics distribution, are presented in the Supplementary Figures S1 and S2, respectively, for comprehensive reference.

Additionally, the performance of the EBM model was compared to an elastic-net-regularized logistic regression, and a gradient-boosting classifier (GBM). Although the performance difference between the EBM and the GBM was negligible, the EBM was selected for its intrinsically interpretable nature. Graphical depictions of the results can be found in the Supplementary Figure S3.

The model was trained using the training set (2018-2020), and the validation set (2021) was used to infer the threshold over the probability trajectories. This threshold allows us to classify patients as “at risk” for developing ovarian cancer at any given time point, considering the model’s monthly prediction. The evaluation is multifaceted, focusing on three key axes evaluated over the test set: (i) machine learning performance metrics, (ii) interpretability of the model’s predictions, and (iii) prediction and learning stability.

### Machine learning model performance

The prediction performance of the model was measured using a two-evaluation axis. First the ROC and Precision-recall curves and the Recall Vs the median days of anticipation of the model were plotted (see Figure 2). Then the performance of the model was reported after establishing the threshold to be considered at risk of developing an OC as per the Methods section (see Table 2).

**Figure 2:**
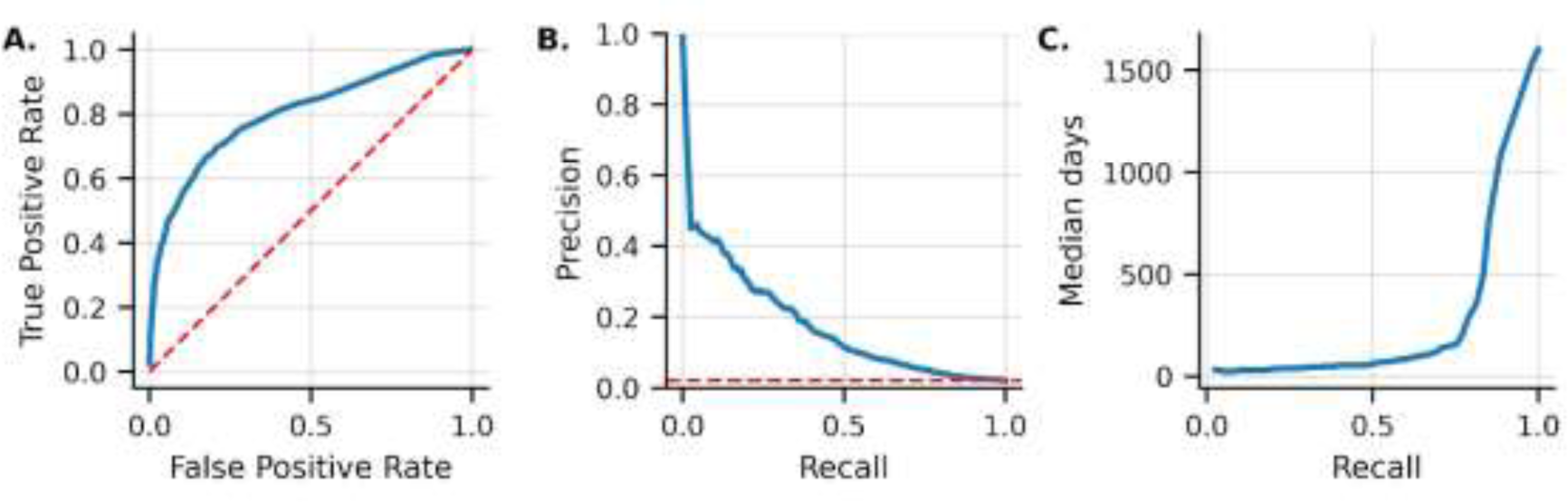
Performance curves on the evaluation set. **A:** ROC-curve, the red dashed line acts as a baseline representing a classifier that makes random guesses; **B:** Precision-Recall curve, the red dashed line is the proportion of the positive class in the set (0.02). This is the precision when classifying all subjects as positive; **C:** recall vs. median number of days between positive prediction and ovarian cancer diagnosis date.

**Table 2:** EBM metrics at a classification threshold of 0.15. Prediction metrics are presented for both the test set (cases diagnosed in 2022 and controls split to the test set) as well as for the test set and all controls (cases diagnosed in 2022 and all controls in our dataset). This is done in order to simulate the continuous evaluation of all women with contact with the public health system. Being, True Positive Ratio TPR= TP/(TP+FN), PPV = TP/(TP+FP), F1-Macro = 2*(Precision * Sensitivity)/(Precision + Sensitivity).

| Metric | Test | Test and all controls |
| --- | --- | --- |
| Sensitivity (TPR) | 0.65 | 0.65 |
| Specificity (1-FPR) | 0.84 | 0.85 |
| Precision (PPV) | 0.07 | 0.02 |
| F1-Macro | 0.52 | 0.48 |
| AUROC | 0.79 | 0.79 |
| AUPRC | 0.16 | 0.06 |
| AUPRGC | 0.96 | 0.98 |
| Positive class prevalence | 0.02 | 0.006 |
| Median days / 365 | 0.28 | 0.28 |

A key aspect of the evaluation involves examining the model’s generalizability and robustness. This was achieved by comparing the performance of the model across the validation and test sets. In particular, Figures 1 and 2 display similar ROC, Precision-Recall, and Recall-Earliness curves for both sets. This consistent performance across datasets suggests good generalizability. In simpler terms, the model effectively transfers its knowledge from the training data to unseen data in the test set, mitigating concerns about overfitting.

Furthermore, the model is resilient to potential data variations that might occur over time due to real-world factors such as the COVID-19 pandemic or changes in how data is collected. This robustness stresses the ability of the model to perform effectively in real-world scenarios.

In conclusion, the consistent performance observed across different years significantly reinforces confidence in the predictions generated by the model and the valuable insights that can be extracted from its results. Moreover, the ability of early detection to generalize and remain robust despite variations in data increases the reliability of the model.

After determining the risk threshold using the probability trajectories computed for the validation set (see Methods section), the performance of the model in decision-making scenarios can be assessed. Table 2 shows the evaluation of the model across two complementary sets, using the metrics described in the Methods section. On the one hand, the scores on the independent test set (2022 data) to assess its generalizability were reported. On the other hand, to simulate continuous monitoring of the whole population under study, metrics obtained by evaluating the monthly probability predictions across the entire study period for ovarian cancer cases diagnosed in 2022 and all control individuals were reported.

As expected, decision-based metrics remain similar in both sets, except for AUPRC and PPV that are extremely influenced by the differences in the prevalence of the positive class. Nevertheless, the AUPRGC (Area Under the Precision-Recall Gain Curve) remains consistent across both sets. This metric specifically addresses the issue of class imbalance by incorporating a weighting scheme, offering a more reliable measure of performance for comparison purposes in such scenarios.

Figure 3 depicts the distribution of prediction lead times for patients in the test set. Although the median lead time is approximately three months (see Table 2), the distribution reveals a relevant aspect: a significant number of patients are predicted to be at risk more than six months in advance of their diagnosis. Notably, some patients are identified as high-risk even a year before their diagnosis.

**Figure 3:**
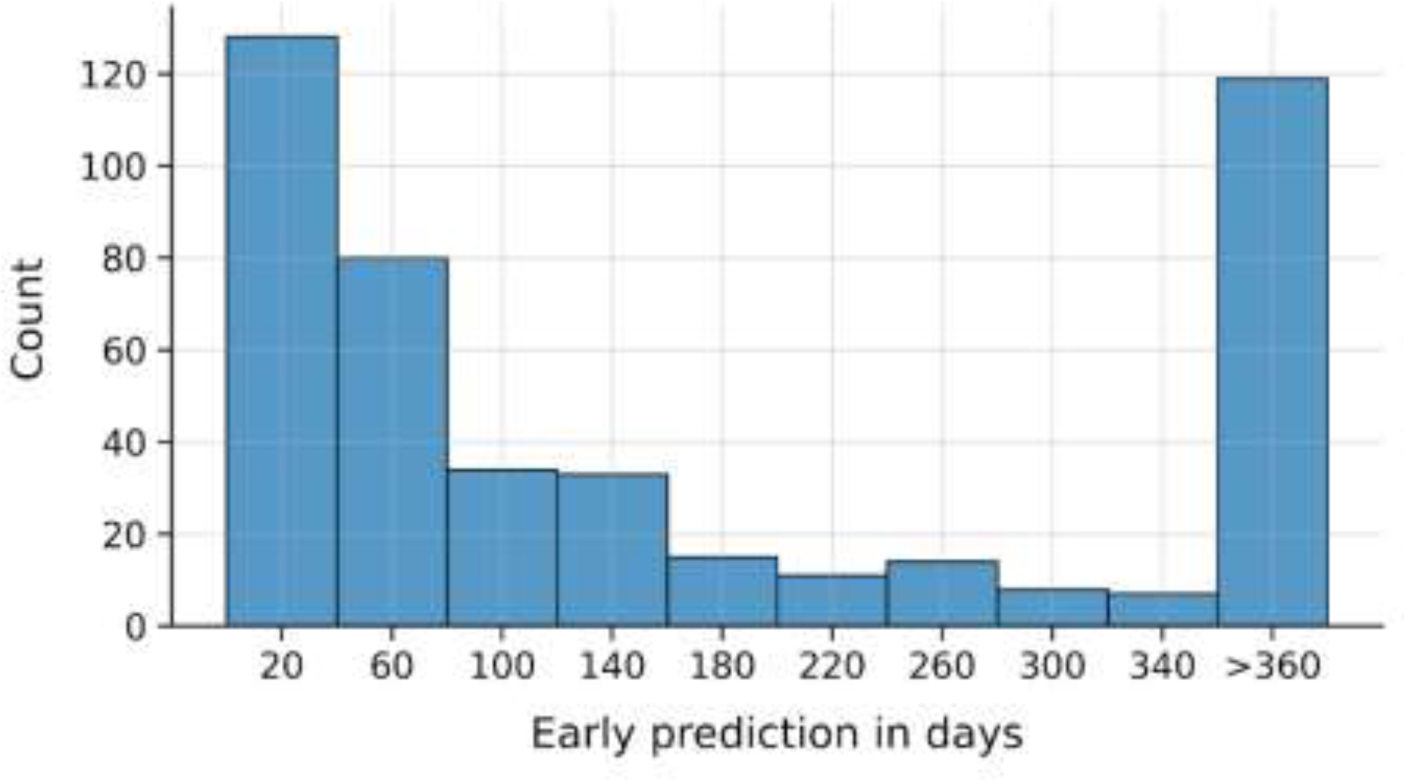
Distribution of the number of days between a case in the test set’s ovarian cancer diagnosis date and a positive classification by our model. Patients with over 360 days between their ovarian cancer diagnosis and their positive classification were grouped.

This early detection rate highlights the potential of the model to flag individuals who might benefit from closer monitoring or earlier intervention strategies.

### Interpretability

Although a comprehensive evaluation of the explainability perspective is beyond the scope of the present work, Figure 4.A shows the most important features influencing the performance of the interpretable machine learning model proposed for predicting ovarian cancer (OC). Longer bars correspond to features that the model considers to be more influential in predicting OC risk.

**Figure 4:**
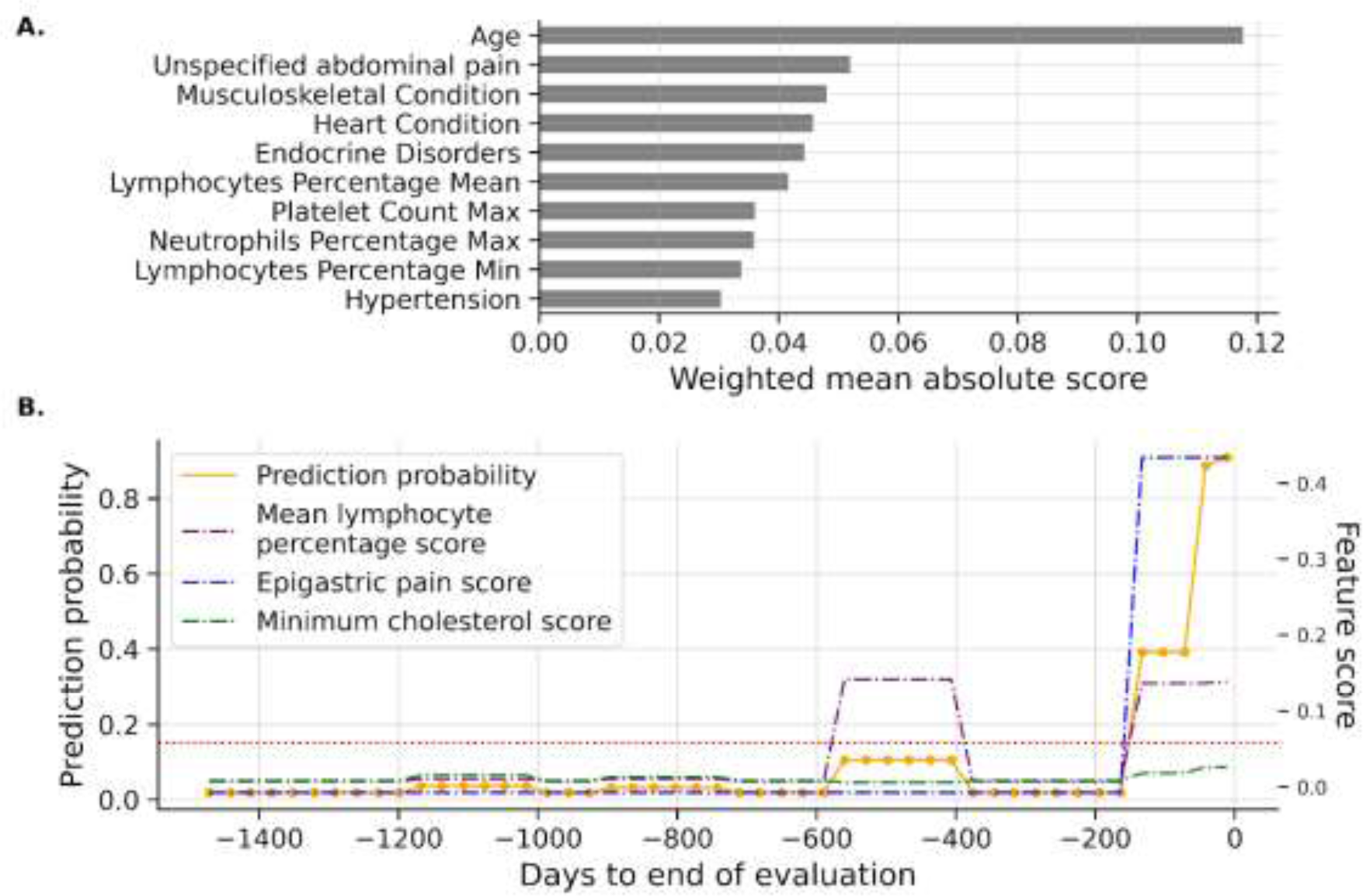
Model interpretation. A: Top 10 weighted mean absolute scores on prediction on the training set. Bar lengths correspond to the relative significance of the feature in the decision-making process of the model. B: Evolution of prediction probability and mean lymphocyte percentage score throughout patient evaluations up until their diagnosis date. The dashed red line is the learned classification threshold.

The most important factors that the model used for assessing the risk include established risk elements such as age, along with other significant features like the existence of abdominal pain, musculoskeletal or heart conditions, and endocrine disorders. Furthermore, the model gives significant importance to certain blood test results, adding information on lymphocyte percentages and platelet counts to its predictions.

Figure 4.B presents key findings regarding the model’s ability to identify patients at risk of developing ovarian cancer (OC). This analysis focuses on two aspects of the model’s performance: early detection potential and interpretable insights. The figure depicts the model’s predicted probabilities of OC for patients over time, visualized as lines representing probability trajectories (orange-like). The x-axis represents the time to OC diagnosis in days, while the y-axis indicates the probability score, ranging from 0 (no predicted risk) to 1 (high predicted risk). These probability trajectories simulate the continuous monitoring of the individuals under study. In this particular case, the probability trajectory of a specific OC patient for four years until her OC diagnosis is shown. The model anticipates the OC diagnosis.

Additionally, the development of the assessment generated by the model can be individually tracked for each patient over time by examining the local explanations of the model, which goes beyond just risk prediction. These interpretability trajectories aim to provide insight into the factors influencing the risk assigned by the model. By analyzing these trajectories with the probability curves, valuable information about how the model classifies patients can be obtained. For example, if a specific risk factor for ovarian cancer (e.g. the history of a particular illness) becomes increasingly prominent in the interpretability trajectory as the probability of ovarian cancer rises, because the model is assigning more weight, it suggests that such risk factor might be more relevant for this particular patient. This interpretability aspect provides detailed information to understand the decision-making process followed by the model and potentially identify key factors that are linked to a heightened risk of ovarian cancer. The example in Figure 4.B shows the interpretability trajectories for Mean Lymphocyte Percentage, Epigastric Pain, and Minimum Cholesterol, in a specific patient. Although the Mean Lymphocyte score increases at - 600 days, it is not enough to classify the patient as at risk. Half a year before the ovarian cancer diagnosis, the model assigns high values to all the mentioned features, which pushes the patient’s ovarian cancer probability beyond the threshold, indicating that the model accurately predicted the onset of ovarian cancer.

Finally, an ablation study was conducted to assess the relevance of each feature group. Feature groups were systematically removed from the dataset, and model fitting and performance were evaluated using the proposed splitting strategy. Supplementary Figure S4 in the supplementary material reveals that the analytics and diagnostics groups are most impactful, as performance significantly decreases when either group is removed.

These results are particularly encouraging as they suggest the model not only predicts OC risk but also offers insights into the factors driving those predictions. This combination of early detection and interpretability has the potential to improve risk assessment and intervention strategies, potentially leading to better patient outcomes.

### Model stability

Figure 5 shows different aspects of the stability in model performance across 100 different random seeds. Figures 5A and B show the distribution of predicted probabilities between a benchmark seed (seed 1) and all other seeds. In an ideal scenario, these sections should be closely distributed around zero, indicating minimal discrepancies in predicted probabilities between the reference seed and other training runs. Figure 5 A and B show that the predictions made by the model for both the control and case groups, respectively, do not exhibit excessive sensitivity to the particular random seed employed. Figure 5C provides an analytical examination of the consistency with which the model attributes significance to different features over several training iterations (one for each seed). The hyperbolic-weighted tau (hwt) statistic was computed for each pair of rankings, subsequently plotting the ensuing distribution. A distribution centered near 1 indicates that the model consistently prioritizes the same features as relevant for ovarian cancer prediction, irrespective of the randomness of the seed employed, which is the case for the model. Finally, Figure 5D provides a comprehensive overview of model performance metrics across all random seeds. These metrics include measures like True Positive Rate (TPR), False Positive Rate (FPR), Positive Predictive Value (PPV), Area Under the ROC Curve (AUROC), Area Under the PR Curve (AUPRC), and the median normalized earliness of ovarian cancer prediction. Consistent values across seeds for these metrics would further solidify the model’s stability and generalizability, suggesting its ability to perform reliably even with slight variations in the training process. In summary, the results suggest that the model exhibits good stability and generalizability, with reliable predictions not significantly influenced by random variations.

**Figure 5:**
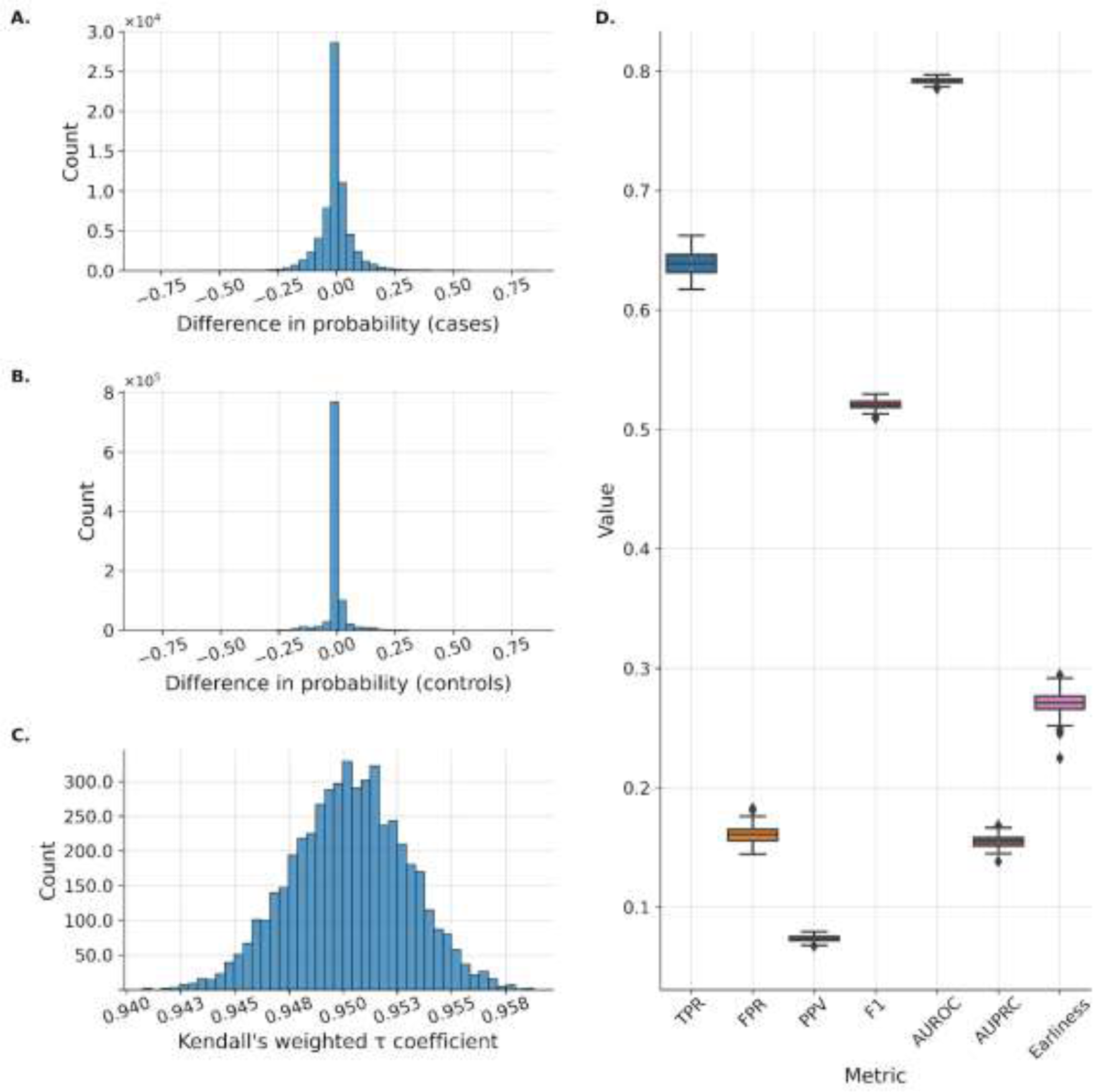
Model stability across 100 different random seeds in model training. A: Count of the difference between the predicted ovarian cancer probability in seed 1 and the predicted ovarian cancer probability in seeds 2-100 in ovarian cancer patients. B: Count of the difference between the predicted ovarian cancer probability in seed 1 and the predicted ovarian cancer probability in seeds 2-100 in non-ovarian cancer patients. C: The stability of the relevance values learned by inspecting the differences across the rankings using the hyperbolic-weighted tau (hwt) statistic. D: True positive rate (TPR), false positive rate (FPR), positive predictive value (PPV), the area under the receiving operating curve (AUROC), the area under the precision-recall curve (AUPRC) and earliness of ovarian cancer prediction as a fraction of a year across random seeds.

## Discussion

This retrospective study of a population of 3,088 patients with an OC diagnosis between 2018 and 2022, as well as 114,942 controls of similar characteristics, used features from 37 regular laboratory tests, 126 symptoms and non-chronic diagnoses, 21 chronic diagnoses, visits to specialists (gynecology and gastroenterology) and age to train a model for accurate identification of patients with ovarian cancer. Despite the model uses only clinical data contained in the patient’s EHR, and does not utilize tumor markers of specific OC tests, it demonstrated consistent and good performance, achieving a sensitivity of 0.65 and a specificity of 0.85.

Some machine learning based approaches for early OC risk have recently been proposed to predict different endpoints related to OC, such as OC type, prognosis or OC risk. A model that used several clinical variables from patients, including blood routine tests, general chemistry data, and also tumor markers was proposed [36], that explores different algorithms such as Random Forest (RF), Gradient Boosting Machine (GBM), and Light Gradient Boosting Machine (LGBM) methods, achieving an accuracy of 88.00%, a sensitivity of 97.00%, and an AUROC of 87.00%. In another study, different combinations of biomarkers associated with OC, where serum samples were obtained from the UKOCST [40] screening trial, were evaluated [37]. The best performance, with an AUC of 0.971 and 0.987 for Bayesian change-point detection algorithm (BCP) and a recurrent neural network (RNN), respectively, was achieved using a combination of Cancer Antigen 125 (CA125) and Human Epididymis protein 4 (HE4). This performance falls to 0.782 AUC with a 47% sensitivity at 90% specificity and 0.8 AUC, and 47.3% sensitivity at 90% specificity when trying to identify ovarian cancer before one year of clinical diagnosis for BCP and RNN respectively. At two years before clinical diagnosis, their best performance was obtained using only CA125 with a sensitivity of 52.7%. When aggregating all the results they reported a median lead time ranging from 1.4 to 1.8 years. A different approach used Support Vector Machine (SVM) and k-nearest neighbors (KNN) algorithms on a dataset containing diverse features from malignant and benign ovarian cancer samples including demographics, blood tests, and ovarian cancer biomarkers [38]. The study reported a mean accuracy over a 10-fold cross-validation of 0.9648 and 0.9724, a specificity of 0.9648 and 0.9724, a precision of 0.9669 and 0.9653 and an F1-Score of 0.9669 and 0.9668 for the SVM and KNN respectively.

Recently, an intricate classifier fusion strategy, based on multi-criteria decision-making (MCDM) for predicting ovarian cancer, was proposed [39]. Initially, 176 base classifiers were created using the top 20 most significant features selected through a feature selection process. Subsequently, a consensus classifier from the 20 highest-performing base models was formed by calculating the weight of each model using MCDM principles. This strategy was applied to a multicenter retrospective-cohort of 10,992 subjects, leading to the creation of two external validation sets: one of them comprising 467 OC and 5,174 control patients, and the other one including 393 OC and 1,951 control patients. The method achieved an AUC of 0.882 with all features, including tumor markers, but this decreased to 0.839 when tumor markers were excluded, with a sensitivity of around 0.65 at a specificity of 0.86. These are similar metrics to those obtained in the approach presented here. However, it must be noted that the authors utilize laboratory tests that, while understandably correlated to ovarian cancer, are not widely tested in the general population without the suspicion of a pathology.

Considering that the model presented here only uses clinical record data found in the general population the performance obtained in predicting early OC risk is quite good, even in comparison to the studies listed in Table 3, which use OC biomarkers. Moreover, many existing studies rely on datasets with significantly smaller sample sizes, which can lead to overfitting and can restrict the generalizability and reliability of their models, potentially leading to inaccurate images of real-world OC prevalence. Furthermore, some studies incorporate specific tumor markers that can increase the precision for early-stage cancer detection but are expensive and unsuitable for large-scale population screening. Finally, factors such as the collection of data in a single center or the failure to consider the temporality of the evaluation process restrict the real-world applicability of several of the studies presented in Table 3.

**Table 3.** Machine learning approaches for OC risk prediction, with their respective sample sizes, accuracy, use of specific OC biomarkers and population used.

| Model | Study size | Total OC | Sensitivity | Specificity | OC markers | Prediction | Population |
| --- | --- | --- | --- | --- | --- | --- | --- |
| Our model | 118,030 | 3,088 | 0.65 | 0.85 | No | OC 1 year | general |
| RF, GBM, LGBM [36] | 349 | 349 | 0.88 | 0.97 | Yes | OC/benign OC | OC patients |
| RNN [37] | 37,293 | 88 | 0.47 | 0.90 | Yes | OC 1 year | OC risk |
| SVN, KNN [38] | 349 | 349 | 0.97 | 0.97 | Yes | OC/benign OC | OC patients |
| MCDM [39] | 10,992 | 860 | 0.65 | 0.86 | Yes | OC/non OC | OC risk |

The approach presented here specifically addresses these limitations by using a large and comprehensive dataset encompassing regionwide data from all public health centers across Andalusia, which enhances the generalizability and reliability of the model. Furthermore, the model relies on readily available and cost-effective features, such as routine blood tests, diagnoses, use of healthcare resources, etc., which are more suitable for population-wide screening strategies. Additionally, techniques to mitigate overfitting risks were implemented, along with the use of a continuous evaluation schema that takes into account the temporality of patient data. By addressing these shared challenges, the model presented here offers a cost-effective, broadly applicable, and generalizable solution for early-stage OC detection.

While the proposed model can be used as an early warning system, it can also be useful as a pre-screening tool within a broader screening strategy. Four key ovarian cancer screening trials have been documented: SCSOCS [41], UKOCST [40], PLCO [32], and UKCTOCS [42]. Unlike these trials, the proposed model does not require extensive screening methodologies or repeated testing, minimizing the need for active patient participation. Direct comparison is complicated by varying reported false positive rates. UKOCST and UKCTOCS only report confirmed cancer cases through surgery, differing from the focus of the proposed model on earlier detection (see Table 4 for a comparison).

**Table 4:** General results of the four main ovarian cancer screening trials conducted thus far. The presented specificity is that obtained in the first line of screening and not at the time of ovarian cancer diagnostic surgery. This specificity was not available for the UKOCST, and UKCTOCS studies, as only false positives at diagnostic surgery were published. Two separate screening methods were employed in the UKCTOCS trial, multimodal screening (MMS) and ultrasound screening (USS).

| **Trial** | **Study size** | **Total cancers** | **Sensitivity** | **First line specificity** | **Years of follow-up** |
| --- | --- | --- | --- | --- | --- |
| Our model | 118,030 | 3,088 | 0.65 | 0.85 | 0 |
| SCSOCS | 41,688 | 35 | 0.77 | 0.91 | <2 |
| UKOCST | 37,293 | 88 | 0.86 | Not provided | <1 |
| PLCO | 34,253 | 126 | 0.58 | 0.9 | <1 |
| UKCTOCS - MMS | 50,084 | 237 | 0.84 | Not provided | <1 |
| UKCTOCS - USS | 50,623 | 221 | 0.73 | Not provided | <1 |

SCSOC and PLCO trials provide data on false positive rates during screening. In the SCSOC trial, 9% of 41,688 patients without ovarian cancer were recommended for medical evaluation or early screening recall. Of these, 77% of ovarian cancers were detected within two years for compliant women. Similarly, in the PLCO trial, 9.7% of 34,253 patients without ovarian cancer were recommended for medical examination, with 66% of cancers detected within one year for compliant women, and 58% when including non-compliant women as false negatives. For UKOCST and UKCTOCS, cancer detection rates at less than one-year follow-up for compliant patients were analyzed. The UKOCST study involved annual transvaginal ultrasound screening, detecting 86% of cancers in compliant patients. The UKCTOCS study evaluated a multimodal screening approach (MMS) and vaginal ultrasound. MMS detected 84% of cancers, while vaginal ultrasound detected 73% in compliant patients within one year. Non-compliance in ovarian cancer screening is a significant limitation, with compliance rates decreasing over time. In the PLCO study, compliance for CA-125 measurement and transvaginal ultrasound decreased from 85% and 84% at first screening to 75% and 73% at later screenings. Similar declines were observed in the UKCTOCS and SCSOC trials, suggesting the need for improved patient compliance to enhance screening sensitivity.

The proposed model shows promise with moderate sensitivity (65%) and fair discrimination (AUROC of 0.79), and its low precision (7%) is mitigated by several advantages. Firstly, the model accurately reflects the low prevalence of ovarian cancer, offering realistic performance expectations. By increasing the sample size, the performance of the model is expected to align closely with real-world scenarios. Secondly, when used as a first-line screening tool, the model provides significant cost benefits due to its minimal upfront costs. Early detection becomes a priority, and even with a false positive rate of 0.15, the model effectively identifies potential cases for further evaluation. This allows specialists to prioritize and see more patients overall, improving the efficiency of the screening process. The simplicity of the model and low patient burden further enhances its practicality and potential for widespread implementation in OC screening strategies.

Finally, another interesting aspect is the interpretability of the model. The use of explainable methods for the derivation of the predictor provides useful information on the most relevant variables used to make decisions. Supplementary Table S4 lists the laboratory test variables used for modeling in the year prior to their index date, ordered by mean absolute relevance score. Although an exhaustive discussion of all the variables is beyond the scope of this manuscript, it is worth commenting on some of the most relevant ones to demonstrate how the model assigns more importance in the classification to OC-related features. The three most relevant analytics variables for the model are lymphocyte, platelet and neutrophil percentages, respectively. Actually, the value of the neutrophil-to-lymphocyte ratio has already been proposed as a potential variable to differentiate between ovarian cancer and benign ovarian disease [43], late-stage and early-stage ovarian cancer [44], and as an indicator of treatment prognosis [45–48]. Interestingly, the neutrophil-to-lymphocyte ratio has shown to even be a valuable indicator of ovarian cancer in cases where the concentration of ovarian cancer antigen 125 (CA125), the main ovarian cancer tumor marker, remains low [49]. Furthermore, an increase in neutrophils [50] and a decrease in lymphocyte count and percentage [46,51], which is observed in Supplementary Table S4, have been directly associated with OC. Also, an increase in the platelet-to-lymphocyte ratio, already observed, has also been proposed as a diagnostic [52,53] and prognostic biomarker [47,54] in ovarian cancer. Furthermore, an increase in platelet counts has been directly associated with OC [55–58]. The eosinophils percentage, which is the eighth most relevant variable, has also been associated with ovarian cancer [43].

In the case of red blood cells, various parameters have been shown to be associated with an ovarian cancer diagnosis at different stages. Hemoglobin concentration, red cell distribution width, and hematocrit have been shown to decrease in ovarian cancer patients as compared to the control cohort [59], with a continued decrease in later ovarian cancer stages as compared to early-stage ovarian cancer [59]. In addition, several blood parameters have also been associated with a poor OC prognosis, such as low red cell distribution width [60], and low hemoglobin concentration [61,62].

On the other hand, dysregulated cholesterol metabolism, as reported in Supplementary Table S4 has been described as a metabolic hallmark in several cancers, including OC [63]. In the case of glucose analytics, differences are expected in the cases with respect to controls due to the Warburg effect in the cancer, reflected as a higher glucose consumption [64]. A similar case, although in the opposite direction, are potassium levels, since EAG K+ channels are proven to be overexpressed in ovarian cancer patients [45].

### Conclusions

A predictor of risk of developing ovarian cancer has been developed, based uniquely on variables contained in the patient’s electronic health records. Different time horizons and historical spans have been considered. As expected, the further the predictive horizon is, the worse the performance of the model is. The predictor achieves a sensitivity of 0.65 and a specificity of 0.85, with an area under the receiving-operating curve (AUROC) of 0.79. To our knowledge, this is the first proposal for an early predictor of ovarian cancer risk based solely on the data registered in the health system from the general population. Although the precision cannot be compared to predictors using molecular data or OC biomarkers, the advantage is that it does not requires prior suspicion of diagnosis and can be automatically used for pre-screening or preventive purposes over hospital data. Therefore, its use to support OC screening is straightforward and inexpensive and can constitute a model exportable to other cancer types.

## Author contributions

VO analyzed and interpreted the patient data, AEM and LA did the data curation and participated in the analysis of patient data, DMM and RV curated the variables and interpreted the data, CL coordinated the analysis of the data and revised the manuscript, JD coordinated the study and wrote the manuscript.

## Acknowledgements

This study was funded by AstraZeneca project “Retrospective observational study for the development of early predictors of high grade serous ovarian cancer” (ES-2021-3211) and is also supported by grants PID2020-117979RB-I00 from the Spanish Ministry of Science and Innovation and grant IE19_259 FPS from Consejeria de Salud y Consumo, Junta de Andalucía.

## Data availability

The datasets analyzed during the current study are available in the Population Health Database repository (https://www.sspa.juntadeandalucia.es/servicioandaluzdesalud/profesionales/sistemas-de-informacion/base-poblacional-de-salud), subject to controlled access according to the regulation for the use of medical data for research in the Andalusian Health System (https://www.sspa.juntadeandalucia.es/servicioandaluzdesalud/sites/default/files/sincfiles/wsas-media-sas_normativa_mediafile/2021/resolucion_conjunta_acceso_a_datos_investigacion_def_20211201%28F%29.pdf).

