## Supplementary Table S1 for "Early prediction of ovarian cancer risk based on real world data"

**Supplementary table S1:** Number and percentage of patients who have experienced symptoms and non-chronic diagnoses used as variables for modeling in the year prior to their index date, ordered by mean absolute score. For cases, index dates are their ovarian cancer diagnosis date and for controls, these are their randomly assigned index date from the array of case diagnosis date. Overall population, case and control numbers and percentages are shown. Statistical comparisons between case and control values are carried out through the Chi-squared test. Multiple comparison is corrected through Bonferroni correction.

| Feature name | Overall | Case | Control | P-Value |
|---|---|---|---|---|
| Total | 118030 | 3088 | 114942 | |
| **Unspecified abdominal pain, n (%)** | 5031 (4.3) | 1008 (32.6) | 4023 (3.5) | <0.001 |
| **Epigastric pain, n (%)** | 3551 (3.0) | 271 (8.8) | 3280 (2.9) | <0.001 |
| **Unspecified ovarian cyst, unspecified side, n (%)** | 274 (0.2) | 178 (5.8) | 96 (0.1) | <0.001 |
| **Intra-abdominal and pelvic swelling, mass or lump, n (%)** | 359 (0.3) | 284 (9.2) | 75 (0.1) | <0.001 |
| **Postmenopausal bleeding, n (%)** | 551 (0.5) | 165 (5.3) | 386 (0.3) | <0.001 |
| **Excessive and frequent menstruation with irregular cycle, n (%)** | 570 (0.5) | 134 (4.3) | 436 (0.4) | <0.001 |
| **Abdominal distension (gaseous), n (%)** | 658 (0.6) | 156 (5.1) | 502 (0.4) | <0.001 |
| **Lower abdominal pain, unspecified, n (%)** | 455 (0.4) | 116 (3.8) | 339 (0.3) | <0.001 |
| **Leiomyoma of uterus, unspecified, n (%)** | 630 (0.5) | 116 (3.8) | 514 (0.4) | <0.001 |
| **Other ascites, n (%)** | 422 (0.4) | 347 (11.2) | 75 (0.1) | <0.001 |
| **Vomiting, unspecified, n (%)** | 1092 (0.9) | 98 (3.2) | 994 (0.9) | <0.001 |
| **Right lower quadrant pain, n (%)** | 166 (0.1) | 45 (1.5) | 121 (0.1) | <0.001 |
| **Unspecified ovarian cyst, right side, n (%)** | 118 (0.1) | 67 (2.2) | 51 (0.0) | <0.001 |
| **Unspecified ovarian cyst, left side, n (%)** | 127 (0.1) | 56 (1.8) | 71 (0.1) | <0.001 |
| **Malignant ascites, n (%)** | 107 (0.1) | 93 (3.0) | 14 (0.0) | <0.001 |
| **Irritable bowel syndrome without diarrhea, n (%)** | 415 (0.4) | 30 (1.0) | 385 (0.3) | <0.001 |
| **Calculus of gallbladder without cholecystitis without obstruction, n (%)** | 1177 (1.0) | 66 (2.1) | 1111 (1.0) | <0.001 |
| **Abnormal uterine and vaginal bleeding, unspecified, n (%)** | 154 (0.1) | 42 (1.4) | 112 (0.1) | <0.001 |
| **Dysphagia, unspecified, n (%)** | 888 (0.8) | 29 (0.9) | 859 (0.7) | 0.266 |
| **Cystocele, unspecified, n (%)** | 404 (0.3) | 34 (1.1) | 370 (0.3) | <0.001 |
| **Acute vaginitis, n (%)** | 737 (0.6) | 38 (1.2) | 699 (0.6) | <0.001 |
| **Change in bowel habit, n (%)** | 144 (0.1) | 14 (0.5) | 130 (0.1) | <0.001 |
| **Benign neoplasm of unspecified ovary, n (%)** | 30 (0.0) | 24 (0.8) | 6 (0.0) | <0.001 |
| **Endometriosis, unspecified, n (%)** | 40 (0.0) | 17 (0.6) | 23 (0.0) | <0.001 |
| **Generalized abdominal pain, n (%)** | 147 (0.1) | 23 (0.7) | 124 (0.1) | <0.001 |
| **Upper abdominal pain, unspecified, n (%)** | 115 (0.1) | 11 (0.4) | 104 (0.1) | <0.001 |
| **Gastritis, unspecified, without bleeding, n (%)** | 528 (0.4) | 48 (1.6) | 480 (0.4) | <0.001 |
| **Complete uterovaginal prolapse, n (%)** | 93 (0.1) | 1 (0.0) | 92 (0.1) | 0.524 |
| **Functional dyspepsia, n (%)** | 267 (0.2) | 16 (0.5) | 251 (0.2) | 0.001 |

| | | | | |
|---|---|---|---|---|
| **Chronic atrophic gastritis without bleeding, n (%)** | 123 (0.1) | 11 (0.4) | 112 (0.1) | <0.001 |
| **Flatulence, n (%)** | 83 (0.1) | 11 (0.4) | 72 (0.1) | <0.001 |
| **Unspecified chronic gastritis without bleeding, n (%)** | 379 (0.3) | 31 (1.0) | 348 (0.3) | <0.001 |
| **Pelvic and perineal pain, n (%)** | 100 (0.1) | 20 (0.6) | 80 (0.1) | <0.001 |
| **Benign neoplasm of right ovary, n (%)** | 44 (0.0) | 32 (1.0) | 12 (0.0) | <0.001 |
| **Left lower quadrant pain, n (%)** | 149 (0.1) | 24 (0.8) | 125 (0.1) | <0.001 |
| **Generalized intra-abdominal and pelvic swelling, mass and lump, n (%)** | 23 (0.0) | 20 (0.6) | 3 (0.0) | <0.001 |
| **Calculus of gallbladder with chronic cholecystitis without obstruction, n (%)** | 63 (0.1) | 1 (0.0) | 62 (0.1) | 1 |
| **Diffuse cystic mastopathy of unspecified breast, n (%)** | 49 (0.0) | 2 (0.1) | 47 (0.0) | 0.368 |
| **Diverticulosis of large intestine without perforation or abscess without bleeding, n (%)** | 999 (0.8) | 69 (2.2) | 930 (0.8) | <0.001 |
| **Calculus of gallbladder and bile duct without cholecystitis without obstruction, n (%)** | 56 (0.0) | 1 (0.0) | 55 (0.0) | 1 |
| **Benign neoplasm of left ovary, n (%)** | 38 (0.0) | 26 (0.8) | 12 (0.0) | <0.001 |
| **Acute vulvitis, n (%)** | 83 (0.1) | 4 (0.1) | 79 (0.1) | 0.173 |
| **Uterovaginal prolapse, unspecified, n (%)** | 220 (0.2) | 12 (0.4) | 208 (0.2) | 0.015 |
| **Subserosal leiomyoma of uterus, n (%)** | 32 (0.0) | 14 (0.5) | 18 (0.0) | <0.001 |
| **Female pelvic inflammatory disease, unspecified, n (%)** | 22 (0.0) | 6 (0.2) | 16 (0.0) | <0.001 |
| **Duodenitis without bleeding, n (%)** | 58 (0.0) | 6 (0.2) | 52 (0.0) | 0.004 |
| **Acute gastritis without bleeding, n (%)** | 70 (0.1) | 5 (0.2) | 65 (0.1) | 0.037 |
| **Esophagitis, unspecified, n (%)** | 141 (0.1) | 8 (0.3) | 133 (0.1) | 0.033 |
| **Female genital prolapse, unspecified, n (%)** | 81 (0.1) | 3 (0.1) | 78 (0.1) | 0.473 |
| **Irregular menstruation, unspecified, n (%)** | 52 (0.0) | 7 (0.2) | 45 (0.0) | <0.001 |
| **Nausea, n (%)** | 419 (0.4) | 30 (1.0) | 389 (0.3) | <0.001 |
| **Vaginal enterocele, n (%)** | 49 (0.0) | 1 (0.0) | 48 (0.0) | 1 |
| **Calculus of bile duct without cholangitis or cholecystitis without obstruction, n (%)** | 518 (0.4) | 30 (1.0) | 488 (0.4) | <0.001 |
| **Crohn's disease, unspecified, without complications, n (%)** | 102 (0.1) | 3 (0.1) | 99 (0.1) | 0.751 |
| **Hemorrhage of anus and rectum, n (%)** | 585 (0.5) | 38 (1.2) | 547 (0.5) | <0.001 |
| **Unspecified acute appendicitis, n (%)** | 36 (0.0) | 2 (0.1) | 34 (0.0) | 0.243 |
| **Rectocele, n (%)** | 134 (0.1) | 5 (0.2) | 129 (0.1) | 0.406 |
| **Colic, n (%)** | 57 (0.0) | 2 (0.1) | 55 (0.0) | 0.662 |
| **Pruritus vulvae, n (%)** | 180 (0.2) | 4 (0.1) | 176 (0.2) | 1 |
| **Other ovarian cyst, left side, n (%)** | 12 (0.0) | 4 (0.1) | 8 (0.0) | <0.001 |
| **Excessive bleeding in the premenopausal period, n (%)** | 50 (0.0) | 9 (0.3) | 41 (0.0) | <0.001 |
| **Calculus of gallbladder with acute cholecystitis without obstruction, n (%)** | 79 (0.1) | 5 (0.2) | 74 (0.1) | 0.056 |

| | | | | |
|---|---|---|---|---|
| **Right upper quadrant pain, n (%)** | 199 (0.2) | 20 (0.6) | 179 (0.2) | <0.001 |
| **Endometriosis of uterus, n (%)** | 11 (0.0) | 4 (0.1) | 7 (0.0) | <0.001 |
| **Heartburn, n (%)** | 240 (0.2) | 7 (0.2) | 233 (0.2) | 0.929 |
| **Acute abdomen, n (%)** | 97 (0.1) | 11 (0.4) | 86 (0.1) | <0.001 |
| **Embryonic cyst of broad ligament, n (%)** | 9 (0.0) | 5 (0.2) | 4 (0.0) | <0.001 |
| **Diverticulosis of small intestine without perforation or abscess without bleeding, n (%)** | 49 (0.0) | 4 (0.1) | 45 (0.0) | 0.039 |
| **Anal fistula, n (%)** | 47 (0.0) | 1 (0.0) | 46 (0.0) | 1 |
| **Diverticulosis of large intestine without perforation or abscess with bleeding, n (%)** | 19 (0.0) | 2 (0.1) | 17 (0.0) | 0.087 |
| **Nausea with vomiting, unspecified, n (%)** | 75 (0.1) | 12 (0.4) | 63 (0.1) | <0.001 |
| **Chronic salpingitis, n (%)** | 15 (0.0) | 5 (0.2) | 10 (0.0) | <0.001 |
| **Diverticulosis of intestine, part unspecified, without perforation or abscess without bleeding, n (%)** | 456 (0.4) | 25 (0.8) | 431 (0.4) | <0.001 |
| **Gastric ulcer, unspecified as acute or chronic, without hemorrhage or perforation, n (%)** | 73 (0.1) | 8 (0.3) | 65 (0.1) | 0.001 |
| **Postmenopausal atrophic vaginitis, n (%)** | 214 (0.2) | 11 (0.4) | 203 (0.2) | 0.036 |
| **Diverticulitis of large intestine without perforation or abscess without bleeding, n (%)** | 44 (0.0) | 3 (0.1) | 41 (0.0) | 0.108 |
| **Chronic superficial gastritis without bleeding, n (%)** | 49 (0.0) | 3 (0.1) | 46 (0.0) | 0.137 |
| **Diaphragmatic hernia without obstruction or gangrene, n (%)** | 1757 (1.5) | 114 (3.7) | 1643 (1.4) | <0.001 |
| **Other intra-abdominal and pelvic swelling, mass and lump, n (%)** | 11 (0.0) | 9 (0.3) | 2 (0.0) | <0.001 |
| **Solitary cyst of unspecified breast, n (%)** | 34 (0.0) | 1 (0.0) | 33 (0.0) | 0.594 |
| **Inflammatory disease of uterus, unspecified, n (%)** | 13 (0.0) | 6 (0.2) | 7 (0.0) | <0.001 |
| **Geographic tongue, n (%)** | 16 (0.0) | 1 (0.0) | 15 (0.0) | 0.346 |
| **Other gastritis without bleeding, n (%)** | 49 (0.0) | 3 (0.1) | 46 (0.0) | 0.137 |
| **Anal fissure, unspecified, n (%)** | 230 (0.2) | 12 (0.4) | 218 (0.2) | 0.023 |
| **Chronic anal fissure, n (%)** | 18 (0.0) | 1 (0.0) | 17 (0.0) | 0.38 |
| **Crohn's disease of both small and large intestine without complications, n (%)** | 13 (0.0) | 2 (0.1) | 11 (0.0) | 0.044 |
| **Chronic or unspecified gastric ulcer with hemorrhage, n (%)** | 8 (0.0) | 1 (0.0) | 7 (0.0) | 0.191 |
| **Female pelvic peritoneal adhesions (postinfective), n (%)** | 12 (0.0) | 6 (0.2) | 6 (0.0) | <0.001 |
| **Unspecified jaundice, n (%)** | 80 (0.1) | 7 (0.2) | 73 (0.1) | 0.005 |
| **Menopausal and female climacteric states, n (%)** | 468 (0.4) | 17 (0.6) | 451 (0.4) | 0.217 |
| **Other ovarian cyst, unspecified side, n (%)** | 14 (0.0) | 3 (0.1) | 11 (0.0) | 0.005 |
| **Left upper quadrant pain, n (%)** | 69 (0.1) | 8 (0.3) | 61 (0.1) | <0.001 |
| **Eosinophilic esophagitis, n (%)** | 5 (0.0) | 1 (0.0) | 4 (0.0) | 0.124 |
| **Other dysphagia, n (%)** | 42 (0.0) | 2 (0.1) | 40 (0.0) | 0.301 |
| **Mild cervical dysplasia, n (%)** | 16 (0.0) | 3 (0.1) | 13 (0.0) | 0.008 |

| | | | | |
|---|---|---|---|---|
| **Acute appendicitis with generalized peritonitis, without abscess, n (%)** | 3 (0.0) | 1 (0.0) | 2 (0.0) | 0.076 |
| **Other noninflammatory disorders of ovary, fallopian tube and broad ligament, n (%)** | 9 (0.0) | 8 (0.3) | 1 (0.0) | <0.001 |
| **Submucous leiomyoma of uterus, n (%)** | 30 (0.0) | 7 (0.2) | 23 (0.0) | <0.001 |
| **Moderate cervical dysplasia, n (%)** | 10 (0.0) | 3 (0.1) | 7 (0.0) | 0.002 |
| **Melena, n (%)** | 213 (0.2) | 13 (0.4) | 200 (0.2) | 0.003 |
| **Anal spasm, n (%)** | 6 (0.0) | 1 (0.0) | 5 (0.0) | 0.147 |
| **Excessive and frequent menstruation with regular cycle, n (%)** | 176 (0.1) | 19 (0.6) | 157 (0.1) | <0.001 |
| **Other cholelithiasis without obstruction, n (%)** | 19 (0.0) | 3 (0.1) | 16 (0.0) | 0.013 |
| **Crohn's disease of small intestine without complications, n (%)** | 20 (0.0) | 2 (0.1) | 18 (0.0) | 0.095 |
| **Endometriosis of ovary, n (%)** | 17 (0.0) | 8 (0.3) | 9 (0.0) | <0.001 |
| **Other impaction of intestine, n (%)** | 48 (0.0) | 2 (0.1) | 46 (0.0) | 0.359 |
| **Intramural leiomyoma of uterus, n (%)** | 56 (0.0) | 17 (0.6) | 39 (0.0) | <0.001 |
| **Dysplasia of cervix uteri, unspecified, n (%)** | 9 (0.0) | 2 (0.1) | 7 (0.0) | 0.022 |
| **Candidiasis of vulva and vagina, n (%)** | 237 (0.2) | 8 (0.3) | 229 (0.2) | 0.597 |
| **Hematemesis, n (%)** | 53 (0.0) | 2 (0.1) | 51 (0.0) | 0.405 |
| **Bilious vomiting, n (%)** | 5 (0.0) | 1 (0.0) | 4 (0.0) | 0.124 |
| **Calculus of gallbladder and bile duct without cholecystitis with obstruction, n (%)** | 18 (0.0) | 1 (0.0) | 17 (0.0) | 0.38 |
| **Diverticulitis of intestine, part unspecified, without perforation or abscess without bleeding, n (%)** | 179 (0.2) | 16 (0.5) | 163 (0.1) | <0.001 |
| **Other ovarian cyst, right side, n (%)** | 13 (0.0) | 3 (0.1) | 10 (0.0) | 0.004 |
| **Diaphragmatic hernia with obstruction, without gangrene, n (%)** | 9 (0.0) | 1 (0.0) | 8 (0.0) | 0.212 |
| **Other specified abnormal uterine and vaginal bleeding, n (%)** | 67 (0.1) | 11 (0.4) | 56 (0.0) | <0.001 |
| **Irritable bowel syndrome with diarrhea, n (%)** | 26 (0.0) | 1 (0.0) | 25 (0.0) | 0.498 |
| **Incomplete uterovaginal prolapse, n (%)** | 53 (0.0) | 1 (0.0) | 52 (0.0) | 1 |
| **Full incontinence of feces, n (%)** | 78 (0.1) | 7 (0.2) | 71 (0.1) | 0.004 |
| **Fecal impaction, n (%)** | 23 (0.0) | 2 (0.1) | 21 (0.0) | 0.121 |
| **Crohn's disease of large intestine without complications, n (%)** | 22 (0.0) | 1 (0.0) | 21 (0.0) | 0.442 |
| **Other esophagitis, n (%)** | 17 (0.0) | 2 (0.1) | 15 (0.0) | 0.072 |
| **Diverticulitis of intestine, part unspecified, with perforation and abscess without bleeding, n (%)** | 7 (0.0) | 1 (0.0) | 6 (0.0) | 0.169 |
| **Other specified inflammation of vagina and vulva, n (%)** | 2 (0.0) | 1 (0.0) | 1 (0.0) | 0.052 |
| **Left lower quadrant abdominal swelling, mass and lump, n (%)** | 2 (0.0) | 1 (0.0) | 1 (0.0) | 0.052 |
| **Chronic duodenal ulcer without hemorrhage or perforation, n (%)** | 3 (0.0) | 2 (0.1) | 1 (0.0) | 0.002 |

| | | | | |
|---|---|---|---|---|
| **Postcoital and contact bleeding, n (%)** | 10 (0.0) | 1 (0.0) | 9 (0.0) | 0.233 |
| **Unspecified foreign body in esophagus causing other injury, initial encounter, n (%)** | 11 (0.0) | 1 (0.0) | 10 (0.0) | 0.253 |
| **Echinococcosis, unspecified, n (%)** | 7 (0.0) | 1 (0.0) | 6 (0.0) | 0.169 |
| **Diverticulosis of intestine, part unspecified, without perforation or abscess with bleeding, n (%)** | 4 (0.0) | 1 (0.0) | 3 (0.0) | 0.101 |
| **Calculus of gallbladder with chronic cholecystitis with obstruction, n (%)** | 2 (0.0) | 1 (0.0) | 1 (0.0) | 0.052 |
| **Acquired atrophy of right ovary, n (%)** | 2 (0.0) | 1 (0.0) | 1 (0.0) | 0.052 |
| **Follicular cyst of left ovary, n (%)** | 3 (0.0) | 1 (0.0) | 2 (0.0) | 0.076 |
| **Periumbilical pain, n (%)** | 12 (0.0) | 2 (0.1) | 10 (0.0) | 0.038 |
| **Calculus of bile duct with acute cholangitis without obstruction, n (%)** | 13 (0.0) | 1 (0.0) | 12 (0.0) | 0.292 |
