## Supplementary Table S2 for "Early prediction of ovarian cancer risk based on real world data"

**Supplementary table S2:** Number and percentage of patients who have been diagnosed with a given group of chronic diseases used as variables for modeling prior to their index date, ordered by mean absolute score. For cases, index dates are their ovarian cancer diagnosis date and for controls, these are their randomly assigned index date from the array of case diagnosis dates. Overall population, case and control numbers and percentages are shown. Statistical comparisons between case and control values are carried out through the Chi-squared test. Multiple comparison is corrected through Bonferroni correction.

| Feature name | Overall | Case | Control | P-Value |
|---|---|---|---|---|
| **Total** | 118030 | 3088 | 114942 | |
| **Musculoskeletal Condition** | 5083 (4.3) | 122 (4.0) | 4961 (4.3) | 0.346 |
| **Heart Condition** | 3917 (3.3) | 85 (2.8) | 3832 (3.3) | 0.084 |
| **Endocrine Disorders** | 5918 (5.0) | 139 (4.5) | 5779 (5.0) | 0.2 |
| **Hypertension** | 6653 (5.6) | 173 (5.6) | 6480 (5.6) | 0.965 |
| **Dementia** | 2334 (2.0) | 28 (0.9) | 2306 (2.0) | <0.001 |
| **Nervous System Disorders** | 3427 (2.9) | 74 (2.4) | 3353 (2.9) | 0.1 |
| **Mental Health** | 4844 (4.1) | 139 (4.5) | 4705 (4.1) | 0.279 |
| **Ischemic Heart Disease** | 3057 (2.6) | 55 (1.8) | 3002 (2.6) | 0.005 |
| **Gynecological Cancers** | 1129 (1.0) | 145 (4.7) | 984 (0.9) | <0.001 |
| **Respiratory Condition** | 4654 (3.9) | 122 (4.0) | 4532 (3.9) | 1 |
| **Digestive System Disorder** | 636 (0.5) | 17 (0.6) | 619 (0.5) | 1 |
| **Breast Cancer** | 3026 (2.6) | 128 (4.1) | 2898 (2.5) | <0.001 |
| **Genitourinary Condition** | 2454 (2.1) | 57 (1.8) | 2397 (2.1) | 0.392 |
| **Rare Cancers** | 1004 (0.9) | 60 (1.9) | 944 (0.8) | <0.001 |
| **Liver Condition** | 3318 (2.8) | 115 (3.7) | 3203 (2.8) | 0.002 |
| **Skin Diseases** | 2296 (1.9) | 68 (2.2) | 2228 (1.9) | 0.327 |
| **Skin Melanoma** | 427 (0.4) | 11 (0.4) | 416 (0.4) | 1 |
| **Respiratory Tract Neoplasms** | 341 (0.3) | 28 (0.9) | 313 (0.3) | <0.001 |
| **Digestive Neoplasms II** | 520 (0.4) | 23 (0.7) | 497 (0.4) | 0.014 |
| **Digestive Neoplasms I** | 1336 (1.1) | 75 (2.4) | 1261 (1.1) | <0.001 |
