## Supplementary Table S3 for "Early prediction of ovarian cancer risk based on real world data"

**Supplementary table S3:** Patient age at index date as well as number and percentage of patients who have had a gynecology or gastroenterology consultation in the year prior to their index date, ordered by mean absolute score. For cases, index dates are their ovarian cancer diagnosis date and for controls, these are their randomly assigned index date from the array of case diagnosis dates. Overall population, case and control numbers and percentages are shown. Statistical comparisons between case and control values are carried out through the Chi-squared test for categorical variables and Mann-Whitney *U* test for continuous variables. Multiple comparison is corrected through Bonferroni correction

| Feature name | Overall | Case | Control | P-Value |
|---|---|---|---|---|
| **Total** | 118030 | 3088 | 114942 | |
| **Age** | 68.0 [59.0, 78.0] | 64.0 [57.0, 74.0] | 68.0 [59.0, 78.0] | <0.001 |
| **Gynecologist visits, n (%)** | 962 (0.8) | 409 (13.2) | 553 (0.5) | <0.001 |
| **Gastroenterologist visits, n (%)** | 4619 (3.9) | 523 (16.9) | 4096 (3.6) | <0.001 |
