## Supplementary Table S4 for "Early prediction of ovarian cancer risk based on real world data"

**Supplementary Table S4:** Median, Q1 and Q3 of analytics values used as variables for modeling in the year prior to their index date, ordered by mean absolute score. For cases, index dates are their ovarian cancer diagnosis date and for controls, these are their randomly assigned index date from the array of case diagnosis date. Overall population, case and control values are shown. Statistical comparisons between case and control values are carried out through the Mann-Whitney *U* test. Multiple comparison is corrected through Bonferroni correction.

| | Units | Overall | Case | Control | P-Value |
|---|---|---|---|---|---|
| **Lymphocytes Percentage, median [Q1,Q3]** | % | 29.9 [21.4,38.3] | 18.3 [9.1,27.9] | 31.0 [22.6,39.0] | <0.001 |
| **Platelet Count, median [Q1,Q3]** | $10^3$U/µL | 233.0 [192.8,278.0] | 276.0 [191.5,387.8] | 231.5 [192.8,275.0] | 0.062 |
| **Neutrophils Percentage, median [Q1,Q3]** | % | 58.0 [49.5,67.3] | 67.9 [61.5,75.0] | 57.2 [49.0,66.1] | <0.001 |
| **HDL Cholesterol, median [Q1,Q3]** | mg/dL | 56.0 [46.0,66.0] | 48.0 [42.8,63.0] | 56.0 [47.0,66.0] | 0.079 |
| **Alanine Transaminase, median [Q1,Q3]** | U/L | 18.0 [13.0,25.0] | 19.0 [13.0,34.5] | 18.0 [13.0,25.0] | 0.358 |
| **Cholesterol, median [Q1,Q3]** | mg/dL | 197.5 [167.0,223.2] | 160.0 [125.5,214.5] | 198.0 [171.0,225.0] | 0.010 |
| **Estimated Glomerular Filtration Rate, median [Q1,Q3]** | mL/min | 73.0 [53.0,86.0] | 80.0 [49.0,87.0] | 72.5 [53.8,86.0] | 0.650 |
| **Eosinophils Percentage, median [Q1,Q3]** | % | 2.1 [1.2,3.3] | 1.6 [0.9,3.1] | 2.1 [1.2,3.3] | 0.159 |
| **Mean Corpuscular Hemoglobin Concentration, median [Q1,Q3]** | g/dL | 32.6 [31.7,33.3] | 32.3 [31.4,33.2] | 32.6 [31.7,33.3] | 0.558 |
| **Basophils Percentage, median [Q1,Q3]** | % | 0.6 [0.4,0.8] | 0.4 [0.3,0.8] | 0.6 [0.4,0.8] | 0.249 |
| **Hemoglobin, median [Q1,Q3]** | g/dL | 13.3 [11.8,14.1] | 11.4 [9.4,12.8] | 13.4 [12.0,14.2] | 0.001 |
| **Creatinine, median [Q1,Q3]** | mg/dL | 0.8 [0.6,0.9] | 0.8 [0.6,1.1] | 0.8 [0.6,0.9] | 0.349 |
| **Urea, median [Q1,Q3]** | mg/dL | 41.0 [32.0,56.0] | 41.0 [27.8,51.0] | 42.0 [33.0,57.0] | 0.468 |
| **Glucose, median [Q1,Q3]** | mg/dL | 98.5 [89.0,124.6] | 113.0 [99.0,131.5] | 97.5 [88.0,123.0] | 0.054 |
| **Monocytes Percentage, median [Q1,Q3]** | % | 7.5 [6.2,9.2] | 7.8 [6.8,8.5] | 7.5 [6.2,9.3] | 0.989 |
| **Potassium, median [Q1,Q3]** | mEq/L | 4.4 [4.1,4.7] | 4.3 [3.7,4.8] | 4.4 [4.1,4.7] | 0.710 |
| **Thyrotropin, median [Q1,Q3]** | µIU/mL | 1.9 [1.2,3.2] | 2.3 [1.8,2.9] | 1.9 [1.2,3.2] | 0.826 |
| **Red Blood Cell Count, median [Q1,Q3]** | $10^6$U/µL | 4.5 [4.1,4.8] | 4.1 [3.1,4.7] | 4.5 [4.1,4.8] | 0.021 |

| | | | | | |
|---|---|---|---|---|---|
| **Mean Corpuscular Volume, median [Q1,Q3]** | fL | 90.2 [86.8,93.3] | 89.8 [80.8,92.8] | 90.2 [86.8,93.3] | 0.441 |
| **Mean Corpuscular Hemoglobin, median [Q1,Q3]** | pg | 29.8 [28.6,30.8] | 29.8 [28.2,30.2] | 29.9 [28.6,30.9] | 0.330 |
| **Total Proteins, median [Q1,Q3]** | g/dL | 6.8 [6.4,7.2] | 6.7 [5.7,7.2] | 6.8 [6.4,7.2] | 0.159 |
| **LDL Cholesterol, median [Q1,Q3]** | mg/dL | 120.5 [88.2,141.8] | 124.0 [105.0,152.0] | 120.0 [86.0,141.0] | 0.524 |
| **Sodium, median [Q1,Q3]** | mEq/L | 141.0 [139.0,143.0] | 140.0 [138.2,141.0] | 141.0 [139.0,143.0] | 0.177 |
| **Hematocrit, median [Q1,Q3]** | % | 39.5 [33.8,42.8] | 34.0 [28.0,39.9] | 39.7 [34.2,42.8] | 0.038 |
| **Phosphorus, median [Q1,Q3]** | mg/dL | 3.5 [3.0,3.9] | 3.1 [2.7,3.8] | 3.5 [3.0,3.9] | 0.145 |
| **Aspartate Transaminase, median [Q1,Q3]** | U/L | 21.0 [17.0,26.5] | 22.5 [17.8,28.5] | 20.0 [17.0,26.0] | 0.448 |
| **Alkaline Phosphatase, median [Q1,Q3]** | U/L | 79.0 [63.0,100.0] | 78.0 [62.2,95.0] | 79.0 [63.0,100.0] | 0.948 |
| **Gamma-Glutamyl Transferase, median [Q1,Q3]** | U/L | 22.0 [15.0,41.5] | 34.5 [21.2,71.2] | 21.0 [15.0,40.0] | 0.045 |
| **Albumin Creatinine Ratio, median [Q1,Q3]** | mg/g | 10.9 [5.4,32.0] | 14.5 [6.4,52.8] | 10.9 [5.4,31.9] | 0.798 |
| **Platelet Distribution Width, median [Q1,Q3]** | % | 13.7 [11.9,16.4] | 11.9 [11.1,14.7] | 13.7 [12.1,16.5] | 0.044 |
| **Creatine Kinase, median [Q1,Q3]** | U/L | 74.0 [50.0,113.0] | 46.5 [37.0,69.5] | 76.5 [52.0,116.2] | 0.002 |
| **Iron, median [Q1,Q3]** | µg/dL | 73.0 [52.0,94.0] | 94.0 [62.0,108.0] | 73.0 [51.2,93.0] | 0.195 |
| **Albumin, median [Q1,Q3]** | g/dL | 4.0 [3.4,4.3] | 3.0 [2.6,3.9] | 4.1 [3.5,4.3] | <0.001 |
| **Chlorine, median [Q1,Q3]** | mEq/L | 103.0 [101.0,106.0] | 103.5 [100.5,105.0] | 103.0 [101.0,106.0] | 0.981 |
| **Large Unstained Cells Percentage, median [Q1,Q3]** | % | 1.7 [1.2,2.2] | 1.3 [0.9,1.8] | 1.7 [1.3,2.2] | 0.031 |
| **Plateletcrit, median [Q1,Q3]** | % | 0.3 [0.2,0.3] | 0.3 [0.2,0.4] | 0.3 [0.2,0.3] | 0.112 |
| **Free Triiodothyronine, median [Q1,Q3]** | pg/mL | 3.0 [2.6,3.3] | 3.2 [2.6,3.3] | 3.0 [2.6,3.3] | 0.786 |
