## Supplementary Figure S1 for "Early prediction of ovarian cancer risk based on real world data"

**Supplementary Figure S1:** Area under the precision-recall curve (AUPRC) of models with different combinations of history and horizon windows.

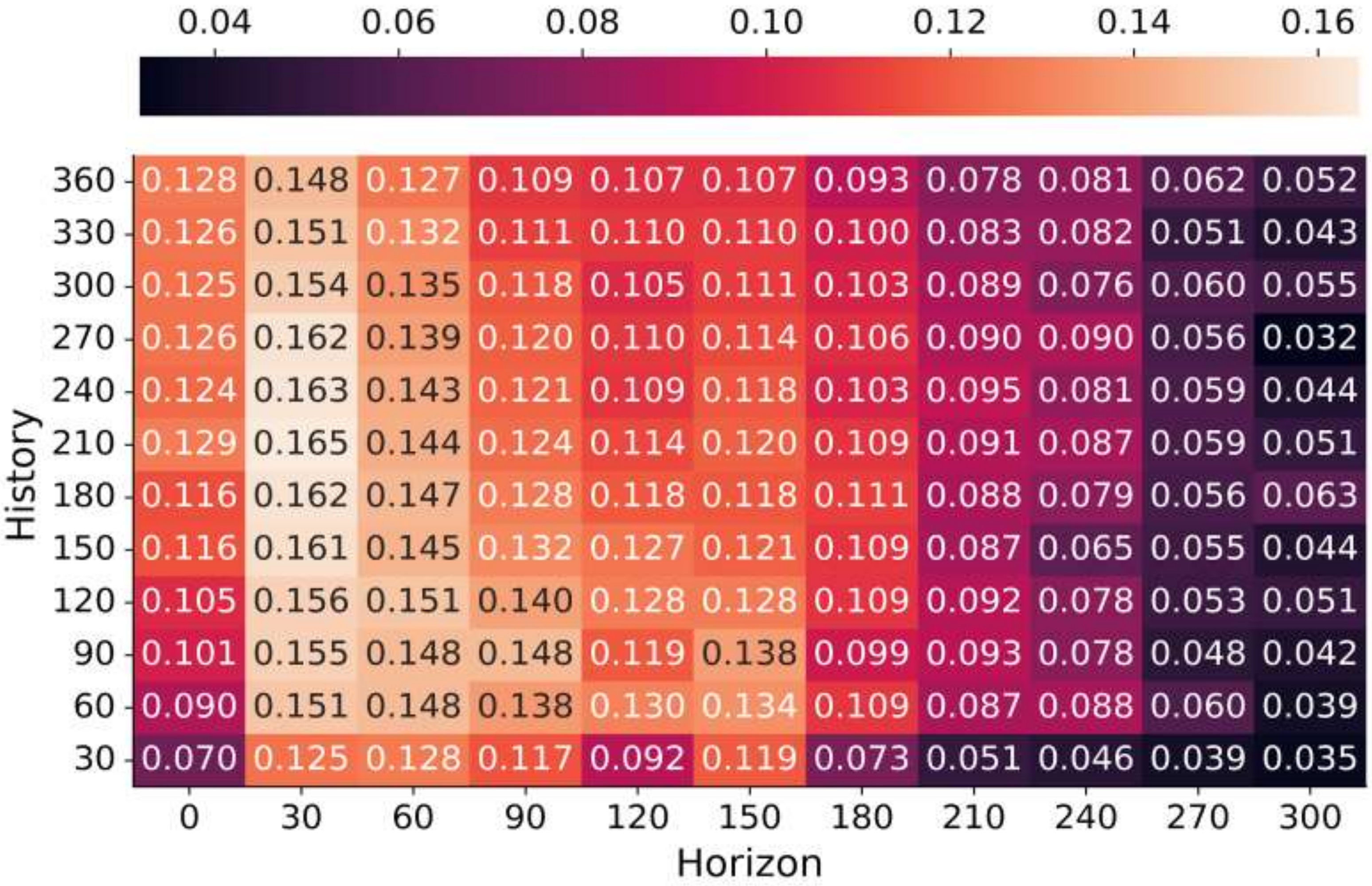
