## Supplementary Figure S2 for "Early prediction of ovarian cancer risk based on real world data"

**Supplementary Figure S2**: AUPR and median earliness in days of models with a 30-horizon window and different history windows across 100 training seeds.

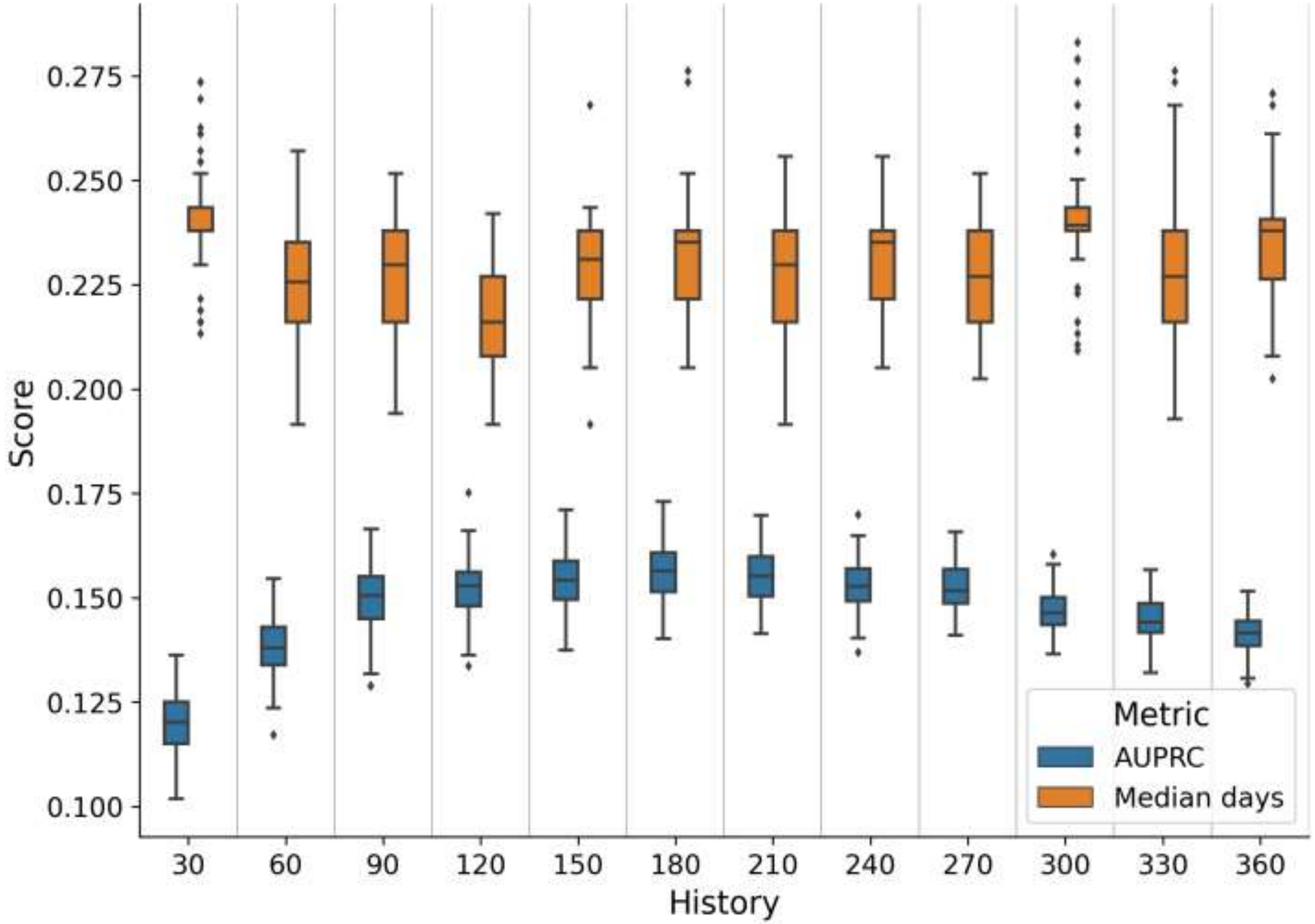
