## Supplementary Figure S3 for "Early prediction of ovarian cancer risk based on real world data"

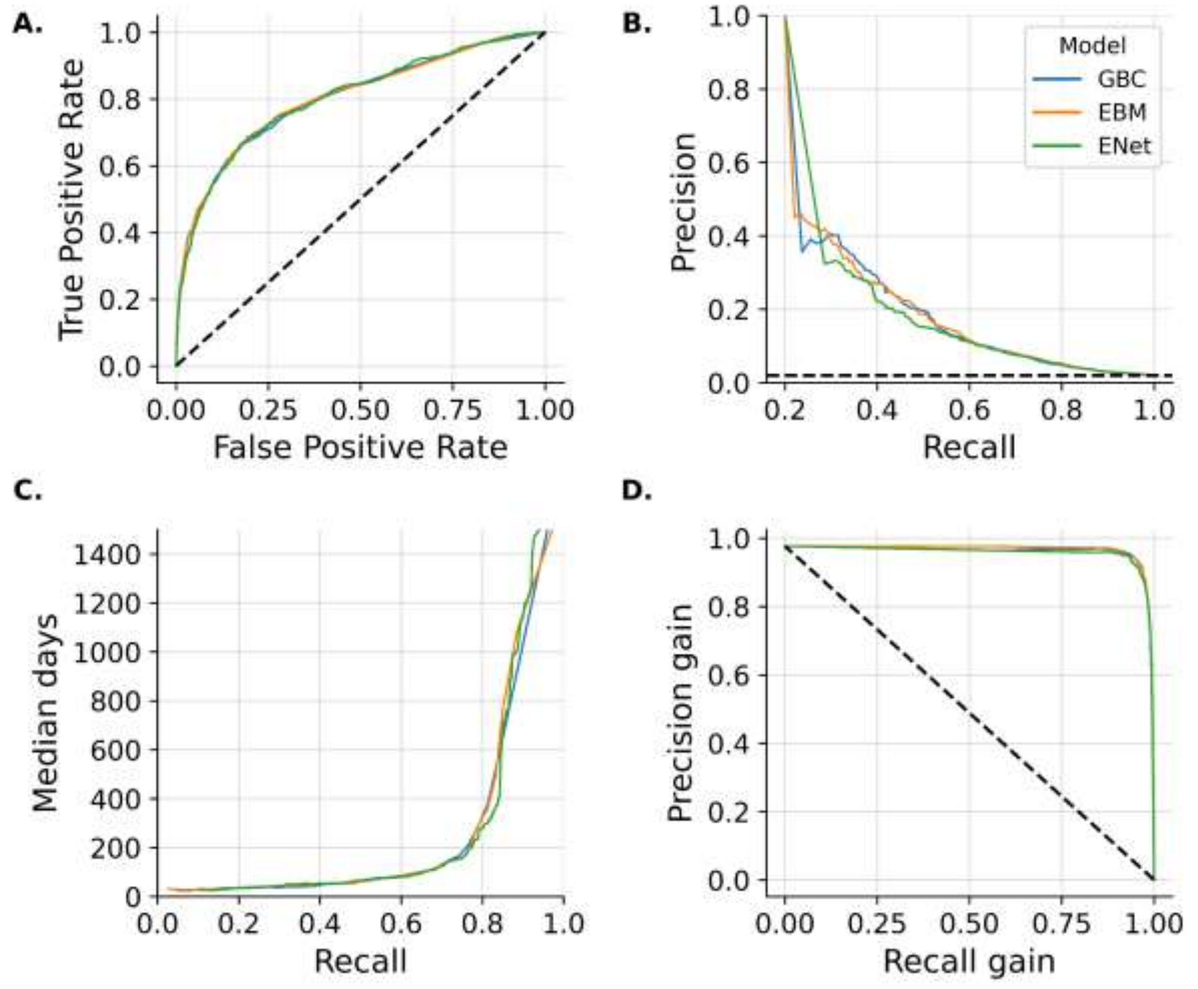


**Supplementary Figure S3:** Comparison of Explainable Boosting Machine (EBM), Gradient Boosting Classifier (GBC) and logistic regression with elastic net regularization (ENet) performance curves on test set. **A:** ROC curve. **B:** Precision-Recall curve. **C:** Earliness in median days-Recall curve. **D:** Precision Gain-Recall gain curve.
