## Supplementary Figure S4 for "Early prediction of ovarian cancer risk based on real world data"

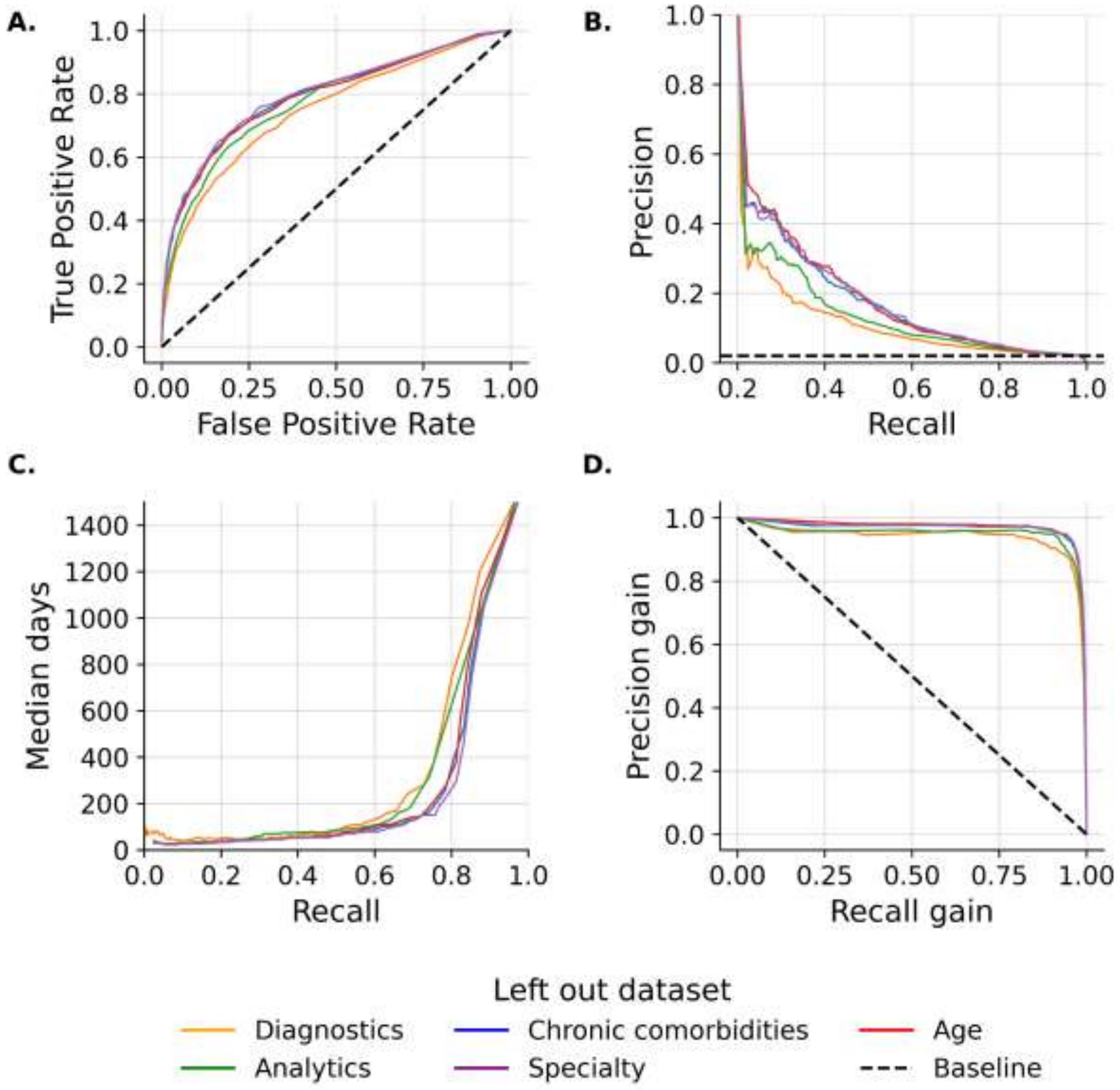


**Supplementary Figure S4:** Ablation analysis investigating the effect of removing different datasets from the predictive model. **A:** ROC curve. **B:** Precision-Recall curve. **C:** Earliness in median days-Recall curve. **D:** Precision gain-Recall gain curve.
